# Circulating Metabolites are Linked to Dementia and Brain Imaging Phenotypes, and Mediate Modifiable Risk Pathways

**DOI:** 10.1101/2025.10.31.25339240

**Authors:** Yue Liu, Dorsa Abdolkarimi, Lachlan Gilchrist, Sara Calhas, Asger Wretlind, Latha Velayudhan, Nicholas J. Ashton, Henrik Zetterberg, Sheena Waters, Cristina Legido-Quigley, Charles Marshall, Petroula Proitsi

**Author notes:** Corresponding author: Corresponding author’s information: Dr. Petroula Proitsi, PhD, Queen Mary University of London, Centre for Preventive Neurology, London, United Kingdom. These authors jointly supervised this work: Cristina Legido-Quigley, Charles Marshall, Petroula Proitsi.

## Abstract

Dementia poses an escalating global health burden, yet its underlying mechanisms remain incompletely understood. In this large-scale, targeted metabolomic study of UK Biobank participants, we applied machine learning models to 327 metabolite and lipid particle measures to identify metabolomic signatures predictive of incident all-cause dementia (ACD), Alzheimer’s disease (AD), and vascular dementia (VaD), beyond conventional risk factors.

Metabolites within these signatures, including the linoleic acid to total fatty acids percentage (LA_pct), glutamine, branched-chain amino acids (BCAAs), low-density lipoprotein (LDL) size, small LDL phospholipids percentage (S_LDL_PL_pct) exhibited widespread associations with dementia outcomes and with neuroimaging markers, including brain atrophy and white matter hyperintensities (WMHs). Many of these key metabolites were associated with plasma neurofilament light chain (NfL) and glial fibrillary acidic protein (GFAP), and most were validated in an independent external cohort. Mediation analyses highlighted that several metabolites potentially mediate the effects of modifiable risk factors—such as obesity, diabetes, hypertension, and education—on dementia risk, with the strongest mediating effects observed for LA_pct in the association between obesity and both ACD and VaD.

Mendelian randomisation (MR) analyses suggested potential causal roles for several metabolites, with the strongest associations being between glutamine and AD and between LA_pct and white matter hyperintensity–related brain atrophy (WMH_atrophy), used as a proxy for VaD. These findings were replicated for glutamine and partially for LA_pct using instrumental variables (IVs) from larger Genome Wide Association studies. The LA_pct associations extended to other polyunsaturated fatty acids (PUFAs), suggesting broader lipid metabolic mechanisms contributing to WMH_atrophy. Statistical colocalisation and expression quantitative trait loci (eQTL) integration revealed shared genetic loci between glutamine, SPRY domain-containing protein 4 (SPRYD4) gene expression levels and AD, and between LA_pct, fatty acid desaturase 1 (FADS1) gene expression levels and WMH-related brain atrophy (WMH_atrophy). Mediation MR further highlighted potentially causal mediating roles for these metabolites in the association between gene expression levels and outcomes. Finally, multivariable MR (MVMR) indicated that glutamine partially mediates the protective relationship between educational attainment and AD.

Overall, most MR associations aligned with neuroimaging-based associations, allowing triangulation of evidence and strengthening causal inference. These findings highlight that blood metabolites -particularly glutamine and LA_pct and other PUFAs-could potentially present as promising biomarkers for early dementia detection and suggest links between modifiable lifestyle factors, metabolic dysfunction, and neurodegeneration, offering potential avenues for targeted prevention in at-risk populations

## Introduction

Dementia represents a growing global public health challenge due to its substantial societal and economic burden ^1^. The insidious onset and progressive nature of dementia often delay diagnosis until irreversible neurodegeneration has occurred, thereby limiting the effectiveness of available therapeutic interventions ^2^. Although new disease-modifying drugs offer clinical benefits in early-stage Alzheimer’s disease (AD)—the most common form of dementia ^3,4^, their effects remain modest, often accompanied by side effects, and typically exclude individuals with co-existing conditions such as cerebrovascular disease. There is therefore an urgent need to better understand the mechanisms underlying dementia and identify novel therapeutic targets and preventive strategies.

Metabolomics—the study of small molecules with a role in metabolism—represents one avenue that may give a deeper insight into the aetiology of different dementias. Blood metabolites sit at the end of the systems biology pathway and represent effective intermediate phenotypes to a given disease because of their proximity to the clinical endpoint ^5^. Metabolites have been reported to interact with amyloid-beta (Aβ) accumulation, driving neuroinflammation, oxidative stress, and tau pathology, contributing to a self-perpetuating cycle of neurodegeneration ^6^. As blood metabolites are potentially modifiable through diet, drugs and lifestyle, and many of them can cross the blood-brain barrier, they can offer a potential window into early neuropathological changes, making them valuable markers of dementia-related biological processes and potential targets for prevention.

While previous studies, including ours ^5,7–10^, have reported associations between blood metabolites and dementia risk, few have systematically investigated how metabolites relate to the future risk of different dementia subtypes, while also accounting for the high intercorrelation among metabolites. Moreover, the relationships between blood metabolites and brain imaging phenotypes—which may reveal early molecular and physiological pathways preceding dementia—remain largely underexplored. Finally, given the close links between blood metabolites and several modifiable dementia risk factors, such as cardiometabolic traits ^11–15^, elucidating the molecular mechanisms through which these risk factors influence dementia risk, offers novel opportunities for prevention, particularly in high-risk populations.

To address these gaps, we performed a large-scale, untargeted metabolomic analysis using data from the UK Biobank to advance our understanding of metabolomic changes associated with incident dementia. Our objectives were: (1) to identify sparse metabolomic signatures associated with incident all-cause dementia (ACD), AD, and vascular dementia (VaD); (2) to assess the associations between key dementia-related metabolites and neuroimaging-derived phenotypes, including global and regional brain volumes, white matter hyperintensities and brain age; and (3) to investigate the potential mediating roles of key metabolites in the relationship between established modifiable risk factors and incident dementia. We employed a triangulation approach, integrating observational analyses, including machine learning models, with causal inference approaches, such Mendelian randomization (MR) and statistical colocalisation, to strengthen the robustness of findings.

## Methods

### Study design and participants

The study was based on data from the UK Biobank study (project ID: 78867), a large-scale prospective cohort study including approximately half a million participants aged 37 to 73 years and recruited from 22 centres across the United Kingdom between 2006 and 2010^16^. All participants provided consent for researchers to access their national health-related hospital and death records. Participant data includes sociodemographic characteristics, lifestyle behaviours, environmental exposures, and health-related factors collected through a combination of a touchscreen questionnaire and nurse-led verbal interviews. The workflow of the study and the exclusion criteria for the participants are summarized in the **Figure 1**.

**Figure 1.**
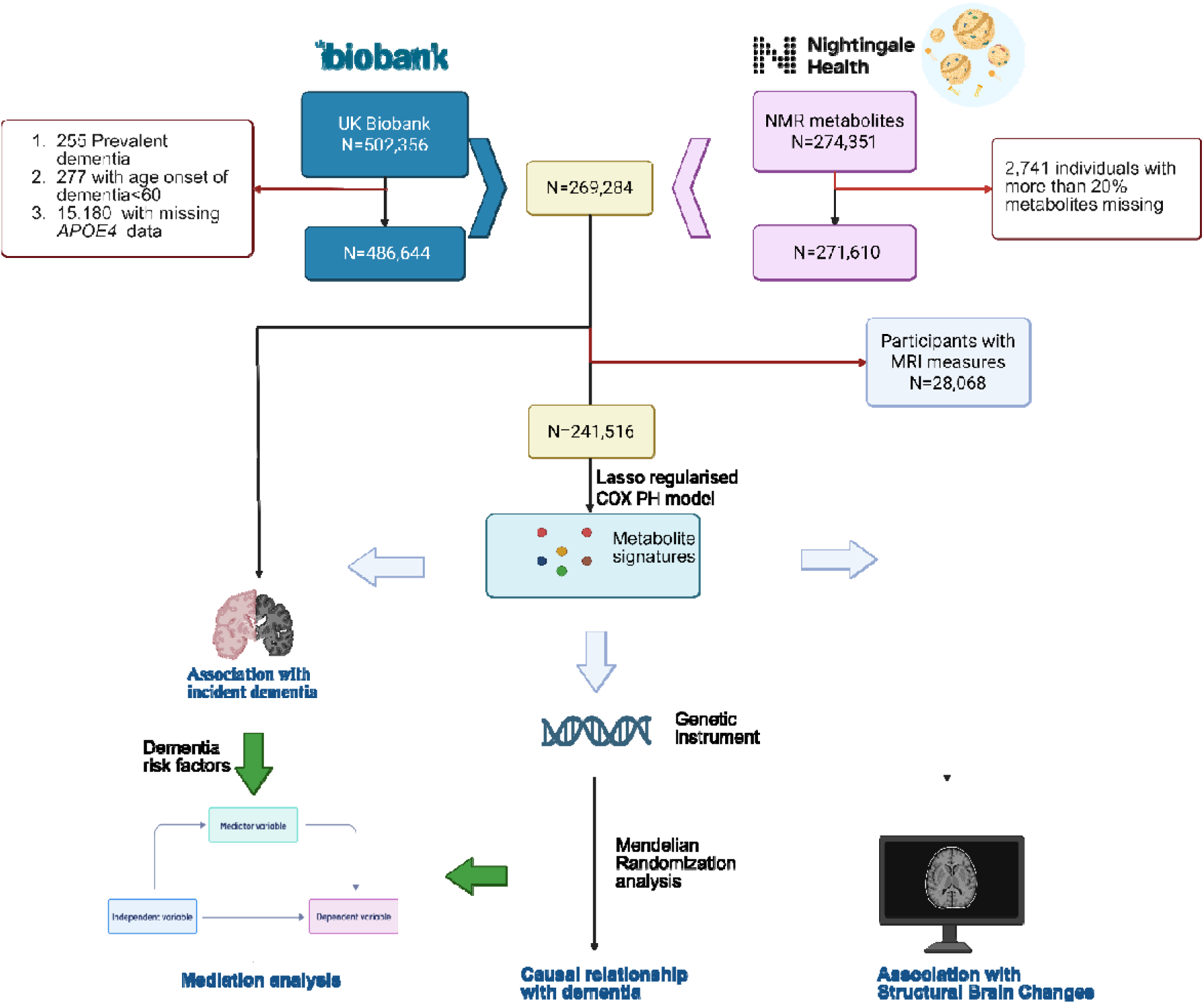
Flowchart. Participants were selected from the UK Biobank and Nightingale Health NMR datasets. Individuals with prevalent dementia, early-onset dementia (age < 60), missing *APOE* ε4 genotype data, or more than 20% missing metabolite values were excluded, resulting in a final sample size of 269,284. Individual Cox proportional hazards (PH) models were used to assess associations between metabolites and incident dementia subtypes: all-cause dementia, Alzheimer’s disease, and vascular dementia. A subsequent LASSO-regularised Cox PH model identified metabolomic signatures and key metabolites associated with each dementia subtype in the non-neuroimaging subset. Key metabolites were further examined for their mediating roles between modifiable risk factors and dementia in the whole dataset, and their associations with neuroimaging phenotypes in a separate imaging subset. Finally, Mendelian randomisation (MR) and multivariable MR analyses were conducted to explore causal relationships between key metabolites and dementia outcomes, as well as their potential causal mediation between modifiable risk factors and dementia.

### Nightingale Metabolomics

Metabolomics were quantified in a random subset of 274,351 UK Biobank participants using nuclear magnetic resonance (NMR) spectroscopy conducted by Nightingale Health Ltd ^17^. The dataset included multiple metabolic pathways and metabolites such as lipids (e.g., cholesterol and fatty acids), amino acids, glycolysis-related metabolites, ketone bodies, and other small-molecule compounds. Detailed information on the assay platform and metabolite quantification is available in the UK Biobank NMR metabolomics companion document (https://biobank.ndph.ox.ac.uk/showcase/ukb/docs/NMR_companion_phase2.pdf).

Metabolite measurements were normalized for known sources of technical variation using the *ukbnmr* R package (https://github.com/sritchie73/ukbnmr/), adjusting a subset of 107 non-derived metabolites for technical variation using robust linear regression and deriving the remaining metabolite measurements post-adjustment. We derived additional metabolite ratios not present in the original Nightingale data but suggested by the *ukbnmr* package authors as having potential biological significance ^18^. In total, 327 metabolite measurements (107 non-derived metabolites, 61 composite metabolites, 81 metabolite ratios, and 76 *ukbnmr*-derived metabolite ratios) were analysed in this study. 271,610 participants were left after excluding participants with more than 20% of missing metabolites. Imputation was performed using the K-nearest neighbour (KNN) clustering method. Metabolite outliers (values > four SDs from the mean) were winsorised. Following imputation and winsorisation, a rank-based inverse-normal transformation was applied to all metabolite measurements to approximate a standard normal distribution prior to performing association analyses. The NMR metabolites list can be found in **Supplementary Table 1**.

### Outcomes

Dementia diagnoses in the UK Biobank dataset were ascertained based on records from hospital inpatient and death registries. Hospital inpatient records were obtained from the Hospital Episode Statistics for England, the Scottish Morbidity Record for Scotland, and the Patient Episode Database for Wales. Death registry records were acquired from the National Health Service Digital for England and Wales as well as the Information and Statistics Division for Scotland. Furthermore, dementia diagnoses were also identified from primary care data utilising disease codes. The diagnoses of dementia were recorded using the tenth edition of the International Classification of Diseases (ICD) coding system. The complete list of codes utilised in this study can be found in **Supplementary Table 2**. Incident dementia events were defined as diagnoses made after the baseline. Participants were censored at the date associated with the incidence of dementia, date of death, or last known follow-up, whichever occurred first. The distribution of time-to-event was illustrated in **Supplementary Figure 1**.

The Imaging-Derived Phenotypes (IDPs) for cortical and subcortical volumes were obtained using FreeSurfer’s aparc atlas (field ID = 192; 66 cortical regions) and ASEG atlas (field ID = 190; 33 subcortical regions). Details of the imaging protocol are available in the open-source document (https://biobank.ndph.ox.ac.uk/showcase/showcase/docs/brain_mri.pdf). Brain- predicted age was generated from raw T1-weighted MRI scans using *brainageR* R package (version 2) ^19^.

### Assessment of covariables and dementia risk factors

The following lifestyle and health-related characteristics associated with dementia risk were included as covariables in all analyses: age at baseline, sex (male/female), ethnicity (White, Asian, Black, Other), recruitment centre (England, Wales, Scotland), and the Townsend Deprivation Index—a measure of material deprivation based on income, employment status, and access to services. The Townsend Index scores range from 0 to 1, with higher scores indicating greater deprivation.

*APOE* ε4 carrier status was determined from genetic data and categorised into 3 groups: non-carriers, carriers of one ε4 allele, and carriers of two ε4 alleles. Additional covariates, included educational attainment (secondary vs. non-secondary education) which is an early-life modifiable dementia risk factor suggested by Lancet commission ^1^ , as well as cardiometabolic risk factors linked to blood metabolites^11–15^: smoking status (never, previous, current), alcohol consumption (never, previous, current), obesity (BMI ≥ 30 kg/m²), and history of diabetes and hypertension. Hypertension was defined as self-reported medication use, prior diagnosis or a mean blood pressure of ≥140/90 mm Hg at baseline assessment. Diabetes at baseline was assessed through self-reports or HbA1c concentrations□≥□6.5% These were derived from self-report questionnaires, hospital episode statistics, and death registry records. Detailed definitions and sources for all covariates are provided in **Supplementary Table 2**.

### Statistical analyses

#### Individual Cox Proportional Hazards Regressions

All investigated outcomes had enough events, meeting the rule-of-thumb requirement of at least 10 events per variable in the model. To address potential inferential bias due to missing covariate values, we performed imputation using KNN method. Cox proportional hazards regression was used to assess the associations between individual blood metabolites and incident dementia outcomes (ACD, VaD and AD). Participants were censored at the date associated with the incidence of dementia, date of death, or last known follow-up. Hazard ratios (HRs) and corresponding 95% confidence intervals (CIs) were calculated. We evaluated the proportional hazards assumption using Schoenfeld residuals. We constructed three models: Model 1 adjusted for age, age^2^, sex, and their interaction terms (age_*_sex, age^2^_*_sex), recruitment centre, ethnicity, and Townsend deprivation index; Model 2 additionally adjusted for *APOE* ε4 carrier status; Model 3 further included modifiable risk factors for dementia (education level, obesity, smoking status, alcohol intake, and histories of diabetes and hypertension).

### Interaction and subgroup analysis

To investigate whether sex and *APOE* ε4 carrier status modified the relationship between metabolites and dementia outcomes, we investigated interactions between metabolites and sex, and metabolites and *APOE* ε4 carrier status, as well as combined metabolites*sex* *APOE* ε4 carrier status interactions. We further conducted subgroup analyses stratifying by sex, *APOE* ε4 carrier status, and their combined categories.

### Lasso-Regularised Cox Proportional Hazards Regression

To develop predictive models, we used the full dataset excluding participants with neuroimaging data. The remaining dataset was then randomly partitioned using the *caret* R package (version 6.0-94) ^20^ into training (50%), validation (25%), and test (25%) subsets, with stratification to preserve the proportion of outcome classes across all splits. Metabolite levels within each subset were inverse rank normalised separately. To identify important metabolites, we applied 10-fold cross-validation with a fixed α of 1 (Lasso regression) to the training data using the *glmnet* R package (Version 4.1-8) ^21^. This process was repeated over 100 iterations. In each iteration, the optimal λ value was selected based on the cross-validation results, and the corresponding regularised coefficients for each metabolite were recorded. Metabolites were then ranked according to the mean absolute value of their regularised coefficients across all iterations, reflecting their overall contribution to the model. To determine the most predictive subset, we evaluated the C-index in the validation set by sequentially adding the top-ranked metabolites in groups of five. The final set of top metabolites was defined as the point at which adding additional metabolites no longer resulted in a statistically significant improvement in the C-index using bootstrapping method. These selected metabolites were used to construct a metabolomic risk score (MetRS) in the test set to evaluate its predictive performance. (**Supplementary Figure 2**).

Incremental improvements in the C-index were evaluated by adding the MetRS to three baseline models (Model 1, Model 2, and Model 3), which were the same as the individual Cox regression models described above. We conducted survival analyses to evaluate the predictive performance of the MetRS for dementia risk at 5-year, 10-year and 15-year intervals. Participants were categorised into a high-risk group (top decile of MetRS) and a reference group (remaining deciles). To flexibly model and visualise the dose**-**response relationship between MetRS and dementia risk, we employed Cox proportional hazards models incorporating restricted cubic splines with three knots.

### Independent replication sample

The AddNeuroMed/Dementia Case Register (ANM/DCR) cohort was used as an independent replication sample to validate the associations of key NMR metabolites with AD and with plasma biomarkers of AD pathology, including neurofilament light (NfL) and glial fibrillary acidic protein (GFAP). Peripheral GFAP and NfL are sensitive markers of neuroinflammation and neuronal damage and are elevated in the preclinical phase of dementia. The ANM/DCR is a multicentre European public**-**private consortium established to facilitate biomarker discovery in Alzheimer’s disease ^22^. AD was diagnosed clinically according to the NINCDS/ADRDA criteria for probable AD. MCI subjects fulfilled the diagnostic criteria for MCI ^23^: (1) memory complaint by patient, family, or physician; (2) normal activities of daily living; (3) mini-mental state examination score (MMSE) range between 24 and 30; (4) geriatric depression scale score less than or equal to 5; (5) clinical dementia rating scale (CDR) score of 0.5; (6) absence of dementia according to the NINCDS-ADRDA criteria for AD. Plasma analyses were performed on an HD-1 analyser (Quanterix, Lexington, MA) at the Department of Psychiatry and Neurochemistry, University of Gothenburg. Commercially available Simoa assays were used to quantify GFAP (GFAP Discovery, #102336). In-house Simoa assays were used to quantify NfL ^24^.

### Associations with Brain Imaging-Derived Phenotypes

Linear regression analyses were conducted in a subset of participants with neuroimaging data (N = 28,068) to examine the associations between key metabolites from each of the three dementia-specific metabolite signatures and neuroimaging markers. We applied three levels of covariate adjustment, consistent with the individual Cox proportional hazards models, with additional adjustment for brain scan centre, intracranial volume and the time interval between baseline and brain scans.

### Mediation analyses

Mediation analyses were performed to investigate whether metabolites identified in LASSO penalised regression models mediate the associations between modifiable risk factors and incident dementia, following the framework by Baron and Kenny ^25^. Linear regression was used to estimate the association between each modifiable risk factor and each metabolite (path a). Cox proportional hazards models were applied to assess the association between each metabolite and incident dementia while adjusting for the corresponding risk factor (path b), and to estimate the direct effect of the risk factor on dementia conditional on the metabolite (path c′). The total effect of each risk factor on dementia (path c) was estimated using unadjusted Cox models. Indirect effects were estimated as the product of the coefficients from paths a and b, and the proportion mediated (prop.Med) was calculated as the ratio of the indirect effect to the total effect, with 95% confidence intervals derived from 1,000 bootstrap samples using the *boot* R package. Indirect effects and prop.Med were calculated only when both path a and path b showed statistically significant associations. Categorical variables, including smoking and alcohol consumption, were dichotomised (0 = never; 1 = former or current) to aid interpretation of mediation estimates. Three models were evaluated in the mediation analysis: Model 1 adjusted for age, sex, recruitment centre, Townsend deprivation index, and ethnicity; Model 2 additionally adjusted for *APOE* ε4 carrier status; and Model 3 further adjusted for all remaining modifiable risk factors.

### Mendelian Randomization (MR) analyses

#### GWAS summary statistics

An overview of the genome-wide association study (GWAS) summary statistics used in this study is provided in the **Supplementary Table 3**. For AD, we utilised summary statistics that excluded proxy phenotyping based on family history from the UK Biobank, given prior evidence that such data can influence effect direction and bias downstream analyses ^26^. Summary statistics for VaD and ACD were obtained from the FinnGen consortium (data freeze 10). The *APOE* locus (chr19:45,020,859**-**45,844,508, GRCh37) was excluded from the dementia GWAS due to its known pleiotropic effects on non-dementia conditions ^27^. For metabolomic traits, we used publicly available summary statistics from a large-scale meta-analysis of nuclear magnetic resonance (NMR)-based GWAS, which included 233 metabolic traits and a combined sample size of 136,016 individuals ^28^.

GWAS summary statistics for dementia risk factors were obtained from established consortia. Specifically, we retrieved data for body mass index (BMI; N = 806,834) from the GIANT consortium ^30^ and educational attainment (EA) summary statistics from the EA4 GWAS meta-analysis (excluding 23andMe) conducted by the Social Science Genetic Association Consortium (SSGAC; N = 765,283) ^31^. We also retrieved summary statistics for obesity from UK Biobank ^32^; systolic and diastolic blood pressure (SBP and DBP) from UK Biobank and the International Consortium of Blood Pressure Genome-Wide Association Studies (ICBP) ^33^; hypertension from a GWAS based on UK Biobank data^34^; smoking and alcohol consumption from a meta-analysis of 60 cohorts ^35^; and diabetes from 32 GWAS conducted in individuals of European ancestry ^36^.

Prior to MR analysis, all summary statistics were QC’d using the *MungeSumstats* R package in R (version 4.2.2). QC was performed with reference to dbSNP build 144 and the *BSgenome.Hsapiens.1000genomes.hs37d5* genome build provided through Bioconductor. The preprocessing pipeline included imputation of missing rsIDs, exclusion of duplicate and multi-allelic variants, and alignment of alleles to the reference genome, with appropriate flipping of effect alleles and frequencies. For datasets provided in GRCh38 coordinates, genomic positions were converted to GRCh37 (hg19) using the UCSC Genome Browser chain file within *MungeSumstats*.

#### Metabolite instrument selection

Independent instrumental variables (IVs) were initially selected based on genome-wide significance (p < 5 × 10□□) using clumping at an r² threshold of 0.001 within a 10,000 kb window around the lead variant. Clumping was performed using the *ieugwasr* R package (v0.1.4), PLINK (v1.9), and the European reference panel from the 1000 Genomes Project Phase 3 (N = 503), restricting to variants with a minor allele frequency (MAF) > 0.01. For exposures with fewer than five IVs at this threshold, a relaxed significance threshold (p < 5 × 10□□) was applied, consistent with previous studies. Metabolites with fewer than five IVs at p < 5 × 10□□ were excluded. When an IV was unavailable in the outcome dataset, a proxy variant in high linkage disequilibrium (r² > 0.8) was identified using LDlink, prioritising proxies with the highest r² and closest genomic distance. Exposure and outcome datasets were harmonized to ensure alignment of effect alleles, and strand-ambiguous palindromic variants with MAF > 0.42 were excluded. Instrument strength was assessed using the F-statistic (F = β²/SE²), and weak instruments (F < 10) were removed ^9^.

#### MR analyses

We conducted bidirectional MR analyses on the casual relationship between key metabolites and dementia using inverse variance weighted MR (IVW-MR) with multiplicative random effects ^37^. We additionally examined WMHs and WMH-related brain atrophy (WMHs_atrophy). WMHs, as the most prevalent form of cerebral small vessel disease, are an established marker of dementia risk. Principal Component 1 (PC1), derived from the shared variance of WMH and cortical thickness via PCA, loads positively on WMH and negatively on cortical thickness, serving as an index of WMHs_atrophy, was included in our study as a proxy of WMHs related VaD ^38^ since the VaD GWA sample size is relatively small. To assess the robustness of primary analyses, several complementary sensitivity analyses were performed using the weighted median, weighted mode, and MR-Egger regression with bootstrapped standard error methods ^39^. The weighted-median approach provides consistent estimates as long as up to 50% of the information is derived from valid IVs^40^. Weighted- mode provides robust estimates when the plurality of similar individual-instrument causal effect estimates originates from valid instrumental variables, even if a significant portion of them is invalid ^41^. We calculated Cochran’s IVW Q-values to identify potential heterogeneity among SNPs included in each analysis. To assess horizontal pleiotropy, we employed MR-Egger regression, wherein the deviation of the Egger intercept from zero was considered significant (P < 0.05) indicative of the presence of horizontal pleiotropy ^42^. We also examined possible pleiotropy using the MR-PRESSO Global test with P values calculated according to 1,000 simulations ^43^. If substantial global heterogeneity was detected, a local outlier test was subsequently conducted to identify any outlier SNPs. Subsequently, the causal effect estimates were reassessed after removing outliers. Metabolites passing sensitivity analyses were further inspected using leave-one-out (LOO) analysis to assess whether causal estimates were driven by the inclusion of a single influential variant. Metabolites that passed all sensitivity criteria were used in subsequent causal mediation MR analysis by performing Multivariable MR (MVMR). Significant MR associations were further validated using metabolite GWAS data from the UK Biobank and the Estonian Biobank ^29^. To avoid sample overlap between GWAS of exposure and outcome, GWAS data from the Estonian Biobank alone were used to replicate associations between metabolites and WMHs and WMHs_atrophy, as these outcome datasets include UKB participants. In contrast, meta-GWAS data were used to replicate associations between metabolites and dementia subtypes.

#### Further analysis of single instrument metabolites

Using LOO analysis, we identified IVs that substantially influenced the IVW-MR estimate when excluded, by rendering the IVW-MR result non-significant. For each of these IVs, we extracted genomic regions ±250 kb and performed statistical colocalisation for each metabolite**-**outcome pair. Regions of interest were harmonised to include only variants present in the reference panel. Colocalisation was performed with the coloc.abf function of the *COLOC* package ^44^ using default priors (1 × 10□□, 1 × 10□□, 1 × 10□□). A posterior probability for hypothesis 4 (PP.H4 ≥ 0.8) was taken as strong evidence of colocalisation, while PP.H4 ≥ 0.6 was considered suggestive. For regions showing colocalisation (including suggestive), we annotated genes corresponding to credible SNPs using the eQTLGen Consortium dataset. For each identified gene, we extracted cis-eQTLs from eQTLGen whole-blood summary statistics within the same ±250 kb window and repeated the colocalisation procedure to evaluate overlap between eQTLs, metabolites, and outcomes. Genes colocalising with both the metabolite and outcome in a given trait pair were then instrumented: independent variants for gene expression were extracted from eQTL summary statistics (p ≤ 5 × 10□□; r² ≤ 0.001; clumping window = 10,000 kb). These were used in IVW-MR to estimate the causal effect of gene expression on both outcome and exposure. When fewer than five SNPs remained after clumping, the wald ratio for the top eQTL was applied. Where gene expression levels showed a significant causal effect on both an outcome and a metabolite, we calculated the proportion mediated using the product of coefficients method as per previous analyses ^45^.

### Causal Mediation Analysis

MVMR was employed to investigate potential causal mediation effects ^5^ and to triangulate the findings from mediation analyses based on observational data. Consistent with the criteria used in observational mediation analyses, evidence of an association between the exposure and mediator (path a), as well as between the mediator and outcome (path b), was first required based on univariable MR analyses. A total causal effect of the exposure on the outcome was not required, as this may be masked by opposing indirect effects via the mediator. We conducted the same MR sensitivity analyses as those performed in the primary MR analyses. MVMR analyses were conducted using the *MVMR* R package ^46^. The indirect effect and proportion mediated were calculated using the delta method to derive confidence intervals.

All statistical tests were two-sided, with p-values < 0.05 considered statistically significant. To account for multiple comparisons, p-values from fully adjusted models were corrected using the false discovery rate (FDR) method.

## Results

### Cohort characteristics

Baseline demographics and health characteristics of participants, stratified by incident dementia status, are summarized in **Supplementary Table 4**. Following the exclusion of individuals with prevalent dementia at baseline, 269,284 participants were included in the analysis including 264,047 controls, 5210 ACD patients, 2311 AD patients and 1213 VaD patients. Compared to the control group, all dementia subtypes exhibited significantly older age, a higher prevalence of *APOE* ε4 carrier status, lower attainment of secondary education, higher Townsend deprivation index scores, increased smoking prevalence, reduced alcohol consumption, and higher rates of hypertension and diabetes. Additionally, ACD and VaD **-** but not AD patients **-** demonstrated a higher proportion of male participants and an increased prevalence of obesity relative to controls.

### Association of individual metabolites with incident dementia

All associations between individual metabolites and incident dementia outcomes are shown in **Supplementary Figure 3** and **Supplementary Table 5**. We additionally investigated associations with several metabolite percentages not previously reported, including the percentage of free cholesterol (FC) and of cholesterol esters (CE) relative to total cholesterol (TC). **For HDL particles,** a higher percentage of FC/TC in large and very large HDL particles was associated with increased dementia risk, while a higher FC/TC in medium HDL particles was inversely associated with dementia risk, particularly in models 1 and 2. **For LDL and VLDL**, most FC/TC in these particles were positively associated with ACD and AD risk, with the exception of small VLDL particles, which showed the opposite effect. Additionally, FC/TC ratios in medium and large LDL particles were negatively associated with VaD.

Overall, we observed that more metabolites were associated with VaD than with AD. However, this number decreased substantially after accounting for modifiable risk factors. (**Supplementary Figure 4 and Supplementary Table 5**).

### Interaction Analysis of Sex, *APOE* **ε**4 Carrier Status, and Individual Metabolites in Relation to Incident Dementia

In interaction analyses, the protective effects of most metabolites on ACD were attenuated both in females and in *APOE* ε4 carriers (*pFDR* < 0.05 for interaction term) (**Supplementary Figure 5 and Supplementary Table 6**). On the other hand, for certain metabolites that were positively associated with ACD — such as total phospholipid proportion, HDL phospholipids, and omega-6 to PUFA ratios—these harmful associations were attenuated in females. We also observed that the protective effect of branched-chain amino acids (BCAAs) appeared to be amplified in *APOE* ε4 carriers, and the detrimental effects of TGs percentages across all lipoprotein types, as well as FC proportions in VLDL and XL-HDL and omega-6 to PUFA ratios, were reduced in *APOE* ε4 carriers. Subgroup analyses supported these interaction patterns.

For AD, the protective effects of cholesterol concentrations or ratios in non-HDL lipoproteins were generally attenuated in *APOE* ε4 carriers **(Supplementary Figure 6 and Supplementary Table 7)**, while BCAAs showed stronger protective in carriers. Subgroup analyses supported these interaction patterns. For VaD, although several metabolites showed interactions with sex and *APOE* ε4, none survived correction for multiple testing (**Supplementary Figure 7 and Supplementary Table**). We finally observed significant positive three-way interactions, indicating that the associations of LA_pct and omega-6 with VaD were modified by both sex and *APOE* ε4 carrier status, with these metabolites showing a markedly stronger protective association with VaD in female *APOE* ε4 carriers compared with female non-carriers.

### Metabolite Signatures for Dementia Prediction

After selecting top metabolites based on incremental improvements in the C-index (**Supplementary Figure 8**), a signature of 15 metabolites predicted ACD and AD, while a signature of 10 metabolites predicted VaD. The metabolomic ACD and AD signatures alone predicted ACD and AD with C-indices of 0.657, 0.655 respectively, while the VaD signature predicted VaD with a C-index of 0.683. The metabolomic signatures significantly improved dementia prediction beyond traditional risk factors including age, sex, *APOE* ε4 carrier status and other modifiable risk factors including education, smoking, alcohol, obesity, diabetes and hypertension. **(Figure 2).** An exception was observed for VaD, where the addition of the MetRS did not statistically enhance predictive performance beyond conventional risk factors. When we looked at how well these MetRS predicted dementia risk over time—specifically at 5-year, 10-year, and 15-year intervals (**Supplementary Figure 9**), the MetRS for ACD and AD demonstrated strong predictive performance within the first 10 years. In contrast, the MetRS for VaD showed meaningful predictive ability starting at 5 years, although this association was not significant after adjusting for modifiable risk factors. We finally observed a dose**-**response relationship between the MetRS and dementia risk, with higher MetRS levels associated with increased risk of dementia (**Supplementary Figure 10**).

**Figure 2.**
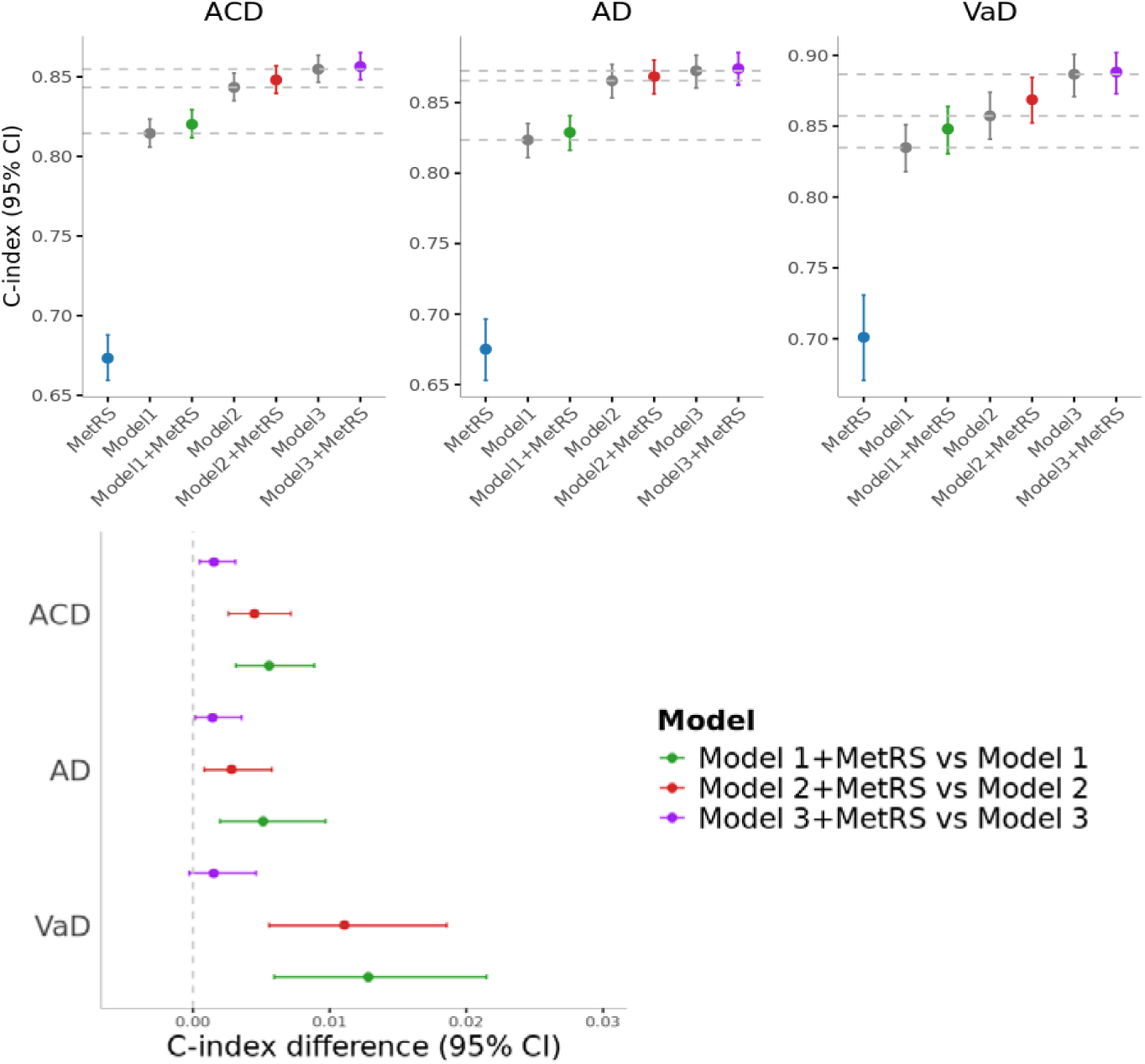
Predictive performance of the metabolomic signatures. Figures (top): Comparison of prediction performance of dementia subtypes (All-cause dementia (ACD), Alzheimer’s Disease (AD), and Vascular dementia (VaD)) using COX PH models trained on the metabolomic risk score only, Age+Sex, *APOE* ε4 status and dementia risk factors, and the sets’ combinations with the metabolomic risk score. Horizontal dashed lines indicate the median performance of the three clinical predictor sets. Figure (bottom): Incremental prediction performance by adding the metabolomic risk score to the model 1 (green), model 2 (red), and model 3 (purple). Model 1 adjusted for age, sex, ethnicity, Townsend index, centre, model 2 additionally adjusted for APOE ε4 carrier status, model 3 additionally adjusted for education level, obesity, smoking, alcohol, history of hypertension and diabetes.

Overall, we observed both overlapping and distinct markers constituting the MetRS for different dementia subtypes (**Figure 3**), with seven of them—leucine, glucose, β- hydroxybutyrate, creatinine, histidine, albumin and LA_pct—being shared across all three subtypes. Each dementia subtype had also unique markers. Triglycerides in large HDL particles (L_HDL_TG), total Lipids in Chylomicrons and Extremely Large VLDL (XXL_VLDL_L), and total BCAA were specific to ACD; citrate and valine were unique to AD; and acetoacetate and LDL size unique to VaD.

**Figure 3.**
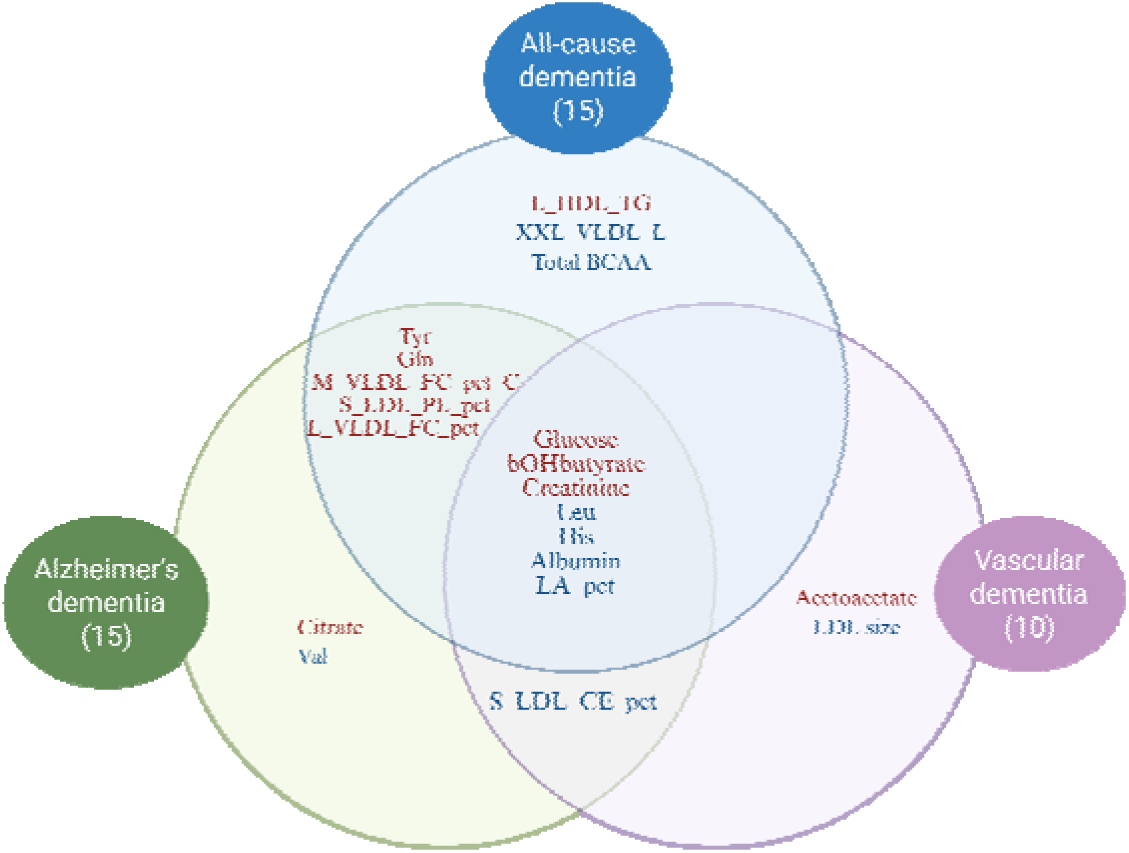
**Venn diagram of metabolomic signatures** of All-cause dementia (ACD), Alzheimer’s disease (AD) and Vascular dementia (VaD). Red color: positive association; Blue color: negative association.

### Validation of Dementia-Related Metabolites in an Independent Cohort

In the independent ANM/DCR cohort, LA_pct showed significant positive associations with both AD and MCI compared to controls (**Supplementary Figure 11**). Glutamine showed a nominally positive and acetoacetate a nominally negative association. We further examined the relationships between these metabolites and AD-related biomarkers (GFAP and NfL) in both the UK Biobank and ANM/DCR cohorts. Notably, glucose, tyrosine, and BCAAs were negatively associated with GFAP or NfL, whereas citrate, creatinine, and LA_pct were positively associated with GFAP or NfL across both cohorts (**Supplementary Figure 11**).

### Association between MetRS, key metabolites and brain imaging phenotypes

Overall, all three MetRSs (ACD, AD, and VaD) were significantly associated with older brain age, reduced brain volumes and enlarged ventricles (**Figure 4**). With regards to regional brain volumes, the VaD MetRS showed the broadest pattern of associations, including volume reductions in the brainstem, cerebellum, and temporal lobes (excluding the medial temporal lobe), greater than those observed for the AD MetRS (**Figure 5**). Univariable regression analyses between key MetRSs metabolites highlighted that two metabolites—LA_pct and glutamine—consistently emerged as protective markers, showing associations with larger brain volumes, reduced ventricular volume, lower WMHs burden, and younger predicted brain age. In contrast, the percentage of free cholesterol in medium VLDL particles (M_VLDL_FC_pct_C) was inversely associated with brain volume and ventricular enlargement (**Supplementary Figure 12**). Regional brain volume analysis showed that LA_pct, glutamine and M_VLDL_FC_pct_C were significantly associated with most brain regions, in line with their associations in global imaging phenotype.

**Figure 4.**
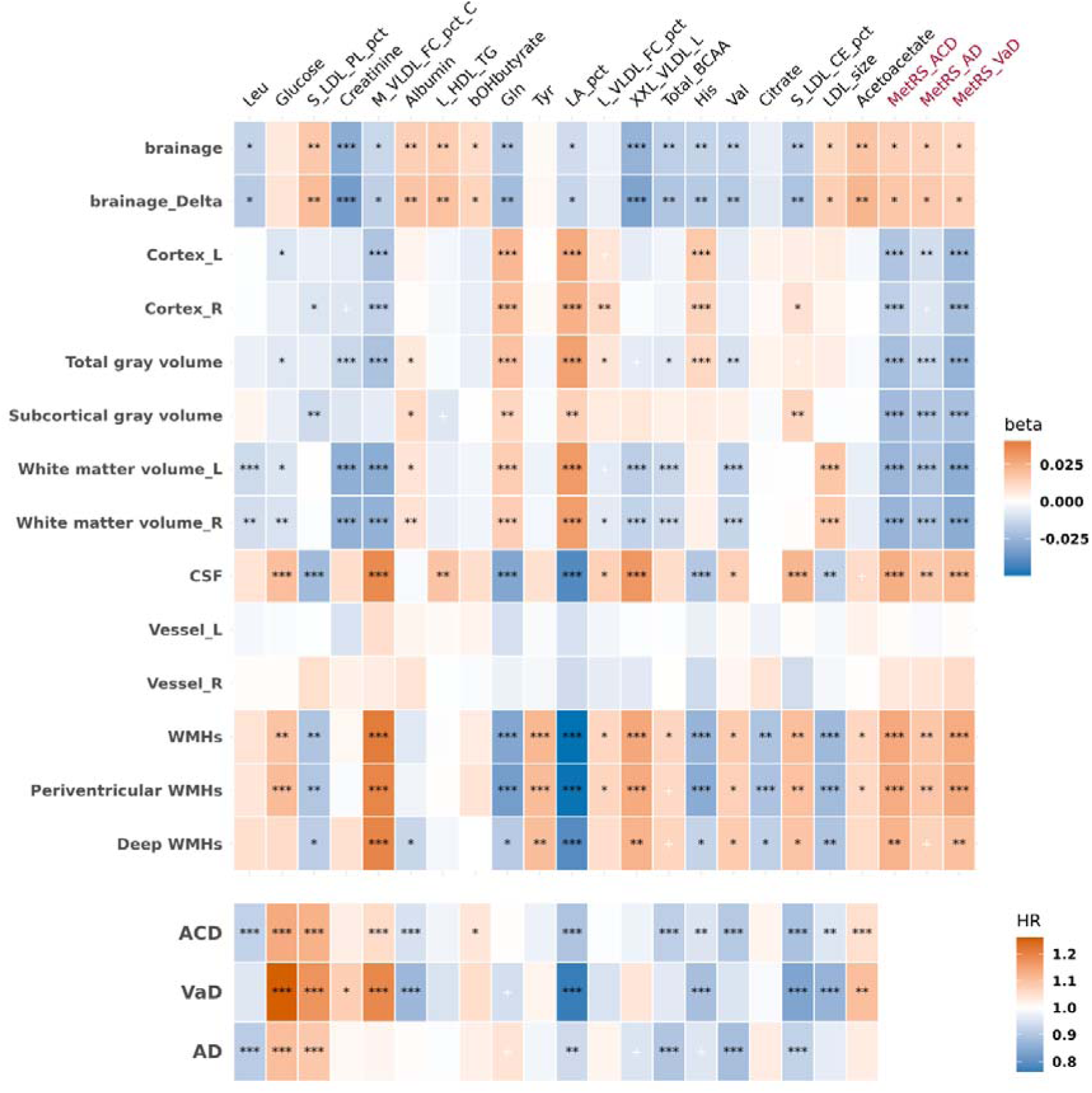
**Associations between metabolomic risk scores (MetRS), key metabolites, and global brain metrics including brain volumes and brain age.** Models were adjusted for age, sex, ethnicity, Townsend deprivation index, assessment centre, and *APOE* ε4 carrier status. ^+^ p<0.05 and FDR>0.05, * FDR<0.05, ** FDR<0.01, ***FDR<0.001

**Figure 5.**
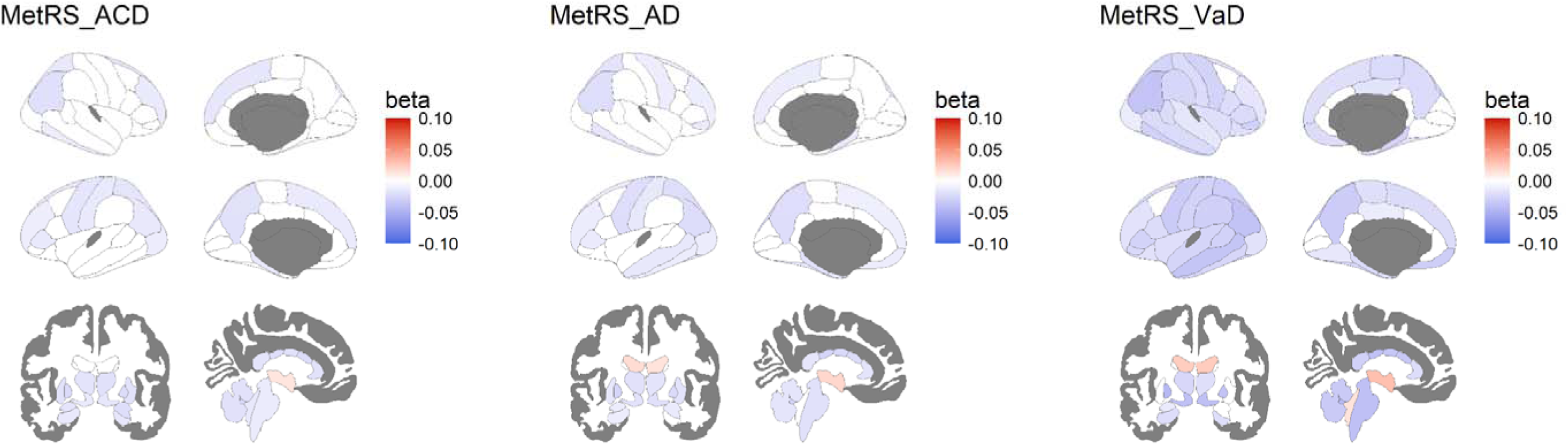
Association between dementia-subtype metabolomic risk scores (MetRS) and regional brain volumes. Dementia subtypes include all-cause dementia (ACD), Alzheimer’s disease (AD), and vascular dementia (VaD). Only statistically significant associations (FDR < 0.05) are displayed. Red regions indicate positive associations, while blue regions indicate negative associations. Non-significant results are shown as blank. Models were adjusted for age, sex, ethnicity, Townsend deprivation index, assessment centre, and *APOE* ε4 carrier status.

### MetRS and Key Metabolites Mediating the Association Between Modifiable Risk Factors and Dementia Risk

The MetRSs for the three dementia types were used to assess their mediating roles in the relationships between modifiable risk factors and dementia types. The associations between education, diabetes, and hypertension and dementia risk were mediated by all three MetRSs (**Supplementary Tables 9 and 10**). Notably, only the VaD MetRS mediated the relationship between obesity and VaD. Among the top mediation effects (prop.Med >30%) observed in model 1 were: Diabetes >> MetRS_AD >> AD, with a proportion mediated (prop.Med) of 34.84% (95% CI: 19.82%**-**83.43%); Obesity >> MetRS_VaD >>VaD, with a prop.Med of 31.91% (95% CI: 18.61%**-**70.26%). These mediating effects were largely preserved even after additional adjustment for *APOE* ε4 carrier status.

When focusing on key MetRS metabolites, several metabolites—particularly glucose and LA_pct—were identified as key mediators of the impact of modifiable risk factors such as obesity, diabetes, hypertension, and education on dementia, with the strongest mediation being observed for the effect of obesity on ACD and AD (**Supplementary Tables 11, 12**, **Figure 6**). For example, glucose and LA_pct mediated over 30% of the association between obesity and both ACD and VaD. However, when additionally adjusting for the remaining risk factors, only glucose and LA_pct mediated over 10% of the effects—specifically, those of obesity and smoking on VaD, and hypertension on AD. We also observed potential suppression effects (defined as indirect effect acting in the opposite direction to total effect) primarily in the relationship between obesity and ACD/VaD, predominantly mediated by BCAAs and percentages of small LDL lipid components (**Supplementary Tables 11, 12, Supplementary Figure 13**). As the association between modifiable risk factors and metabolites were explored cross-sectionally, we also examined the reverse direction— modifiable risk factors mediating the associations between metabolite levels and dementia. These analyses showed stronger and more significant mediating effects (**Supplementary Table 11, 12**). For example, we found evidence that diabetes strongly mediated the association between higher glucose levels and all three types of dementia: (AD: prop.Med = 35.51%, 95% CI: 20.88%**-**73.65%), (VaD: 31.34%, 95% CI: 23.04%**-**43.57%), and (ACD: 28.75%, 95% CI: 19.45%**-**47.11%).

**Figure 6.**
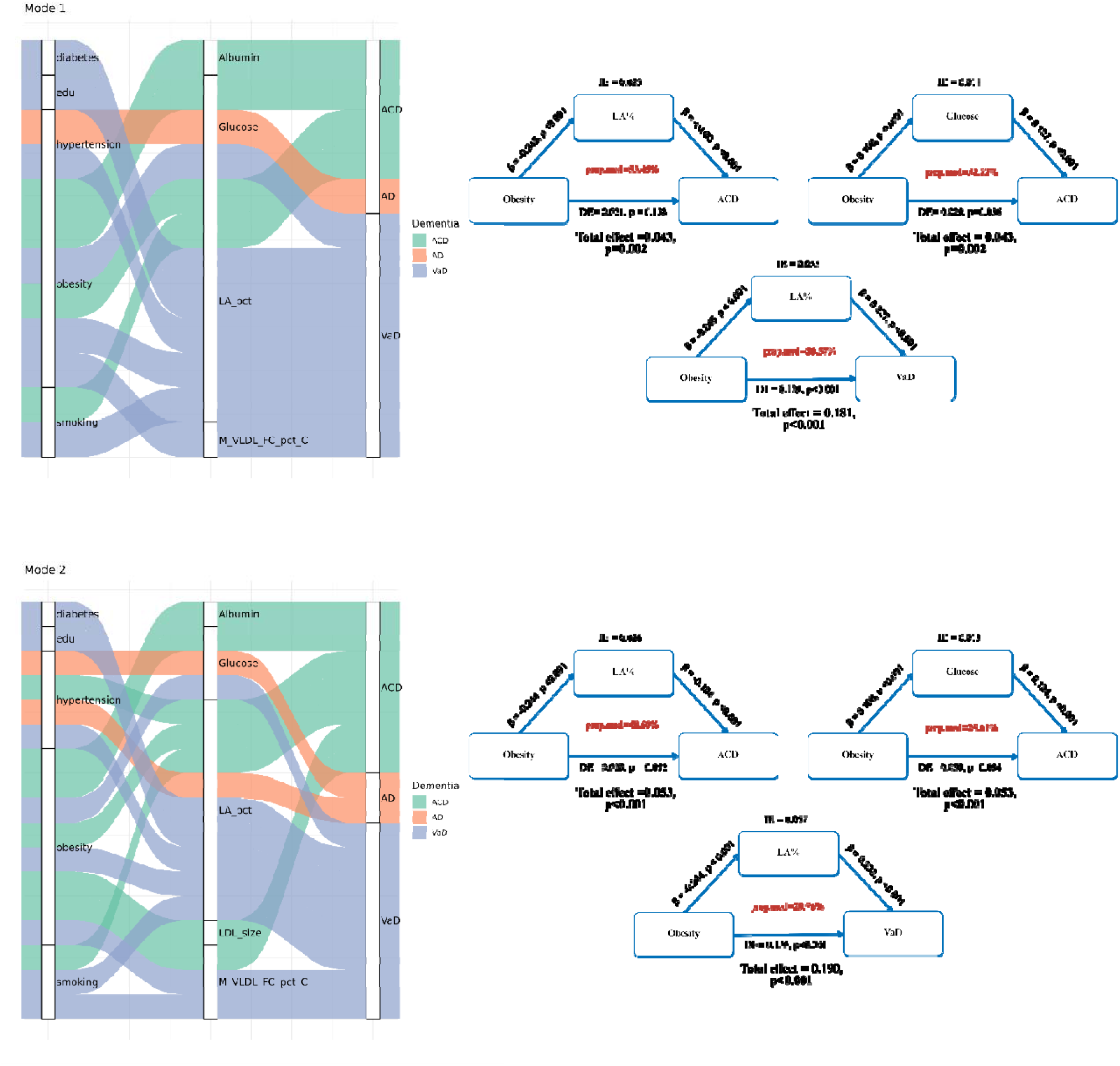
**Mediating effects of metabolomic signatures in the relationship between modifiable risk factors and dementia subtypes:** all-cause dementia (ACD), Alzheimer’s disease (AD), and vascular dementia (VaD). **Left**: Each diagram displays pathways from modifiable risk factors (left panel), through key metabolites (middle panel), to dementia outcomes (right panel), only those with proportion of mediation (Prop.med) >10% are shown in diagrams. **Right:** Mediation path diagram highlighting the strongest effects (Prop.med >30%). Model 1: adjusted for age, sex, ethnicity, Townsend deprivation index and assessment centre; Model 2 additionally adjusted for *APOE* ε4 carrier status.

### MR analyses

We first used MR analyses to assess the bidirectional causal relationships between key MetRS metabolites and dementia subtypes, as well as with key neuroimaging phenotypes (WMHs and WMHs related atrophy) used as VaD proxies. Six associations were found significant (*pFDR*<0.1) (**Supplementary Table 13, Figure 7)**. Overall, the strongest association and the only significant at p*FDR*<0.05 was observed for glutamine, which showed a significant inverse association with the risk of AD (β = **-**0.168, SE = 0.035, *pFDR* < 0.001). Other metabolites associated with AD included S_LDL_PL_pct and leucine (*pFDR*<0.1). We also observed a positive causal link between LA_pct and valine with WMHs-related atrophy. Finally, in the reverse direction, WMHs were causally associated with increased LDL size. All MR associations showed consistent directional effects in sensitivity analyses (MR-Egger, Weighted Median and Weighted Mode) **(Supplementary Table 14)**; however, only the associations of glutamine and leucine with AD, and of LA_pct with WMHs were statistically significant using all three sensitivity methods. Scatter plots of exposure**-**outcome tests are shown in **Supplementary Figure 14**. No pleiotropy (MR-Egger intercept P□>0.05) was observed and although MR_PRESSO indicated pleiotropy in the S_LDL_PL_pct-AD relationship, no outlier SNPs were detected and there was no significant change in distortion (p>0.05 for both tests) (**Supplementary Table 15**). We aimed to replicate the significant (p<0.05) MR associations using metabolite GWAS data from the UK Biobank and the Estonian Biobank. Glutamine was significantly associated with a lower risk of AD (β = **-**0.112, p = 0.004), and LA_pct showed a trend toward a positive association with WMHs_atrophy (β = 0.065, p = 0.081) (**Supplementary Table 16**).

**Figure 7.**
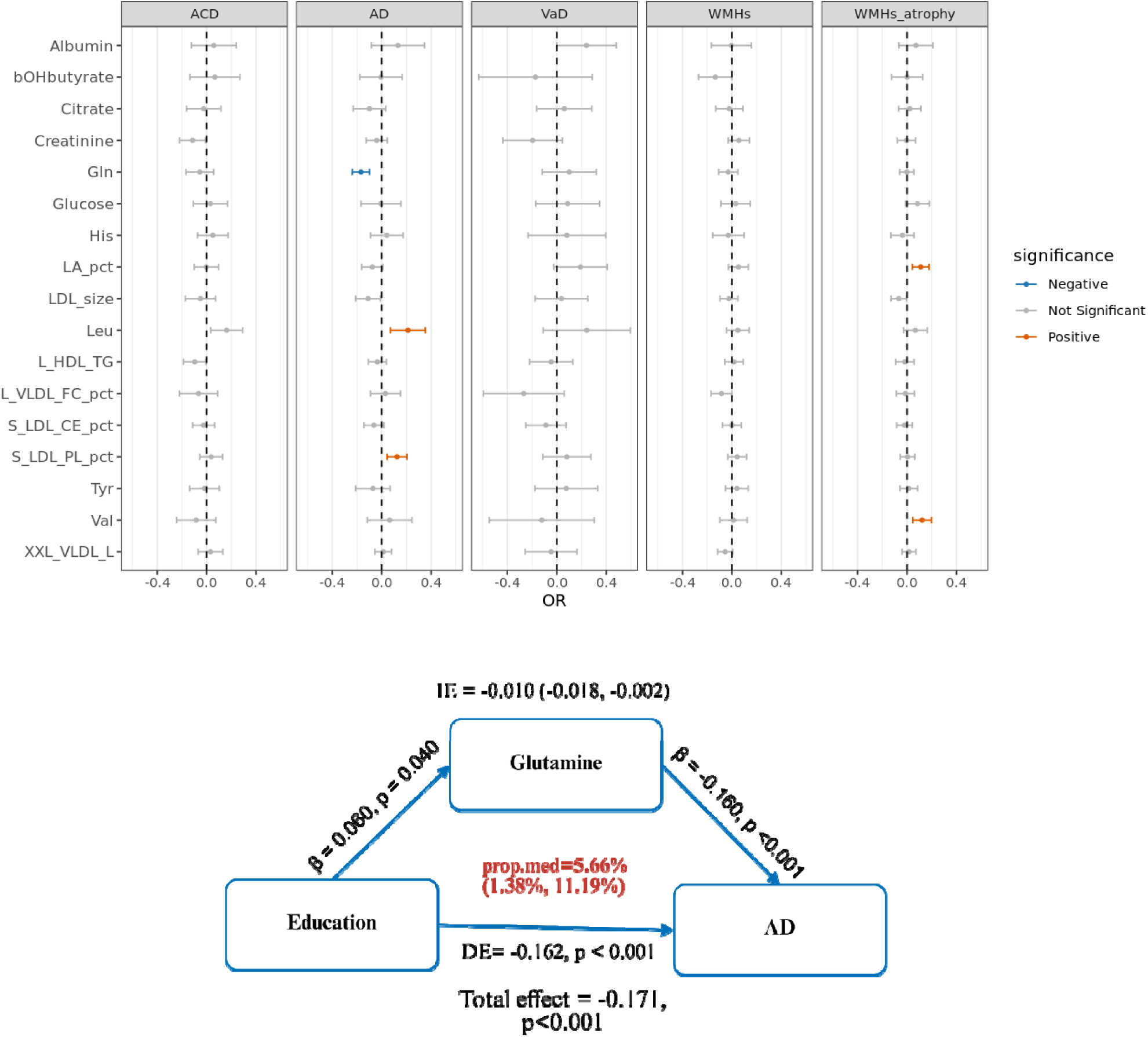
Mendelian randomisation (MR) and mediation MR. **Top:** Causal associations between key metabolites and dementia and related outcomes: all-cause dementia (ACD), Alzheimer’s disease (AD), and vascular dementia (VaD), white matter hyperintensities (WMHs), WMHs related brain atrophy (WMHs_atrophy). **Bottom:** Mediation path diagram showing significant causal mediating effects of key metabolites in the relationships between risk factors and dementia.

LOO analyses identified one SNP with a substantial influence on the combined effect estimates for LA_pct (**Supplementary Table 17**). Removal of rs174528, located upstream of the FADS1 gene in the FADS 1/2/3 cluster, resulted in complete attenuation of the causal effect of LA_pct on WMHs_atrophy, indicating that the observed association was largely driven by this individual variant. Subsequent colocalisation analyses on the +/- 250kb region of rs174528 (Chr11:61293499-61793499) showed strong evidence of colocalisation between LA_pct and WMHs_atrophy (**Supplementary Table 18)**. Further statistical colocalisation analyses were conducted between blood eQTLs at 6 genes at the chr11: 61293499-61793499 region, LA_pct and WHM_atrophy (**Supplementary Table 19)**. Only eQTLs for fatty acid desaturase 1 (*FADS1*) gene expression levels showed statistical colocalisation with LA_pct (PP.H4□=□0.95) and with WMH_atrophy (PP.H4□=□0.84) (**Supplementary Figure 15)**. Suggestive colocalisation was observed between *FADS2* and *TMEM258* with WMHs_atrophy (PP.H4□=□0.76 and 0.65, respectively). MR analyses highlighted that higher *FADS1* expression was causally associated with increased LA_pct (β = 0.51, 95% CI: 0.255**-**0.765, p < 1 × 10□²□□) (Wald ratio; using rs174528 as IV) and with WMHs_atrophy (β = 0.856, 95% CI: 0.813**-**0.899, p = 1.21 × 10□□) (Wald ratio; using rs174528 as IV). Causal mediation analysis subsequently showed that LA_pct partially mediated the effect of *FADS1* on WMHs_atrophy (proportion mediated [95% CI] = 6.63%, 1.41**-**11.86%).

In addition, based on our previous work that highlighted rs2939302 as an influential SNP ^9^ and a very recent AD GWAS study ^47^ reporting a novel AD SNP - rs2657879- which is in perfect LD with rs2939302 (R^2^=1, https://ldlink.nih.gov/), we further investigated whether exclusion of rs2939302 attenuated the causal effect of glutamine on AD. LOO analysis highlighted that removal of rs2939302 resulted in a 17.7% change in the beta estimate, highlighting partial attenuation of the causal effect of glutamine on AD (**Supplementary Table 17**). Subsequent colocalisation analyses (+/- 250kb region of rs2939302 (Chr12:56610020-57110020)) showed strong evidence of colocalisation between glutamine and AD (PP.H_4_□≥□0.8) (**Supplementary Table 18**). Further statistical colocalisation analyses were conducted between blood eQTL for 5 genes at the chr12: 56610020-57110020 region, glutamine and AD (**Supplementary Table 19**). Only SPRY domain-containing protein 4 (*SPRYD4*) showed statistical colocalization with glutamine (PP.H4□=□0.97) and with AD (PP.H4□=□0.82) (**Supplementary Figure 16)**. MR analyses further showed that *SPRYD4* expression showed positive causal associations with glutamine (β = -0.279, 95% CI: -0.159 **-** -0.399, p = 5.64 × 10□□) and with AD (β = 0.088, 95% CI: 0.023**-**0.153, p = 0.007) and that glutamine causally mediated the effect of blood *SPRYD4* expression on AD (proportion mediated [95% CI] = 53.20%, 3.22**-**103.17%).

### MR mediation analyses between metabolites, modifiable risk factors and dementia

To complement observational analyses and strengthen causal inference, we additionally conducted mediation MR analyses to explore whether key causal metabolites mediate the effect of risk factors (RF) on dementia (RF >>metabolites >> dementia) or conversely, whether RFs mediate the effect of key metabolites on dementia (metabolites >> RF >> dementia), using a causal mediation framework. The causal framework included the following pathways: RF>>metabolites, RF>>dementia, and metabolites>>RF. Results are displayed in **Supplementary Table 20.** We identified 65 significant associations (*pFDR* < 0.1), all of which remained robust after outlier correction using MR-PRESSO. LOO did not find single influential SNPs (**Supplementary Tables 21-23**).

For RF>>dementia, we found that greater educational attainment was causally associated with decreased AD risk, whereas higher DBP, SBP, and hypertension were significantly associated increased risk of VaD (**Supplementary Table 23**). No significant associations were observed between modifiable risk factors and ACD. For RF >> metabolites, educational attainment and BMI were associated with glutamine levels, while both diabetes and BMI were associated with leucine levels. For metabolites>>RFs, glucose, histidine, leucine, and XXL_VLDL_L levels were associated with SBP, DBP and hypertension (**Supplementary Table 24**). Subsequent causal mediation analyses showed that glutamine significantly mediated the association between education and AD (**Figure 7, Supplementary Table 24**). When looking at mediation effects of metabolites >> RF >> dementia, hypertension and elevated blood pressure significantly mediated the effects of several metabolites—histidine and leucine—on the risk of VaD, despite the absence of a direct causal association between these metabolites and VaD (**Supplementary Table 24**).

## Discussion

In this large-scale, systematic analysis, we identified sparse plasma metabolomic signatures that predicted incident dementia—specifically AD, VaD and ACD—and which improved risk stratification beyond established clinical risk factors. Several key metabolites were strongly associated with brain volumetric measures and substantially mediated the effects of modifiable risk factors on dementia outcomes. Causal inference analyses highlighted that a subset of metabolites showed evidence of causal associations with AD and WMH**-**related atrophy, used as a proxy of VaD ^38^, with glutamine additionally partially mediating the effect of educational attainment on AD risk. By integrating results from both observational analyses and MR, our study provides evidence implicating early metabolic dysregulation in the pathogenesis of dementia, with potentially important implications for risk stratification and targeted prevention strategies.

LA_pct—identified as a shared protective metabolite across all three MetRS—was consistently associated with larger brain volumes, lower WMH atrophy, and lower future dementia risk; and appeared to mediate the effect of modifiable risk factors, particularly obesity, on dementia in our observational analyses using the UKB. LA is an essential nutrient since it cannot be synthesised in the body. As the most abundant omega-6 fatty acids, it plays a fundamental role in maintaining cell membrane integrity, which is crucial for healthy neuronal function ^48^. Although this pattern was not fully supported by cross-sectional associations in the ANM/DCR cohort, and when looking at the positive association between LA_pct and NfL and GFAP levels, it is consistent with our previous finding that higher LA_pct relates to better cognitive function in late midlife (NSHD)^49^ and with broader epidemiological evidence linking higher LA levels to improved cardiometabolic health, including lower cardiovascular disease and type 2 diabetes risk^50^.

Conversely, our MR analyses suggested that higher LA_pct levels were causally linked to greater WMHs_atrophy, but that this association is driven by genetic variation at the FADS1/2/3 locus. Statistical colocalisation, blood eQTL integration and MR-based mediation further suggested that genetically predicted LA_pct levels (instrumented by rs174528) partially mediate the effect of blood *FADS1* gene expression on WMHs_atrophy. The *FADS1/2/3* cluster is a pleiotropic locus, and SNPs in this region **-** such as rs174528 **-** are known to play a critical role in the regulation numerous polyunsaturated fatty acids (PUFAs) -and other lipid fractions- and are responsible for the desaturation of omega-6 PUFAs in the LA to AA pathway, as well as omega-3 PUFAs in the alpha-linolenic to docosahexaenoic acid pathway ^51^. Our previous MR analyses on the causal effects of metabolites on neuropsychiatric traits using the Metabolon platform also suggested that higher levels of LA-containing lipids were linked to increased risk of depression, anxiety, bipolar disorder and schizophrenia, whilst Arachidonic Acid (AA)-containing lipids had overall protective effects ^9^; a finding also observed in an independent study on bipolar disorder, suggesting that the conversion of LA into AA by the FADS1/2/3 cluster genes may play a key role in the observed associations and ultimately in brain pathology ^51^. AA was not measured in the Nightingale platform, but we examined other PUFAs, with both omega_3_pct and DHA_3_pct showing significantly negative associations with WMHs_atrophy. LOO analyses suggested that these associations also appear to be driven by genetic variation at the FADS1/2/3 locus and specifically rs174528, but in the opposite direction compared to LA_pct (**Supplementary Figure 17**). These findings further support the involvement of the FADS1/2/3 cluster gene variation in PUFA metabolism, brain health and neurodegeneration.

Although liver is the main organ of PUFA metabolism, the *FADS1/2/3* cluster genes are expressed widely across human tissues and cell types, including in the brain ^52^, and genetic variation at the FADS1/2/3 locus has been associated with several brain imaging-derived phenotypes ^53,54^. As PUFAs can also be transported into the brain across the blood-brain barrier it is therefore possible that LA_pct and overall PUFAs can impact white matter atrophy via both peripheral and brain specific mechanisms. However, although variants at the FADS1/2/3 locus are biologically credible instruments because of their direct role in PUFA biosynthesis, their pleiotropic nature means they may violate core MR assumptions and may not be credible instruments^55^. Nevertheless, the statistical colocalisation between *FADS1* expression, LA_pct and WMH_atrophy implicates altered PUFA metabolism as a mechanism linking genetic variation at this locus to WMHs_atrophy.

Overall, our findings highlight that genetic variation in fatty acid metabolism may have a detrimental effect on white matter integrity, at least partly through altered LA_pct and other PUFAs, while higher circulating LA_pct — potentially reflecting a healthier diet — is associated with healthier brain structure and lower dementia risk. Together, these results position LA_pct and related PUFA pathways as key mediators linking genetic risk, diet, modifiable risk factors and dementia-related brain changes, highlighting their potential both for prevention and for therapeutic targeting. Further work is urgently needed to disentangle these effects.

Glutamine levels showed the strongest evidence for a protective causal association with risk of AD in our MR analyses, and this result was further validated using IVs from a larger GWAS. This finding is consistent with our previous MR studies using smaller GWAS datasets for glutamine and AD ^5^, as well as with our recent work where glutamine was measured using the Metabolon platform ^9^. Our cross-sectional analyses with imaging phenotypes also showed that higher glutamine levels were associated with favourable brain structural measures, suggesting a potential direct neuroprotective effect, in agreement with our MR results. Our observational analysis using the UKB produced more mixed results, showing that elevated blood glutamine levels were nominally associated with a decreased risk for incident VaD and decreased NfL levels-in agreement with the MR results- but on the other hand they were also nominally associated with increased risk of AD — the latter finding being validated using the independent ANM/DCR dataset. These contradictory results with MR may reflect reverse causation or residual confounding common in observational analyses. Further MR sensitivity analyses using leave-one-out MR, showed that excluding rs2939302 located on 12q13.3, near the Glutaminase 2 gene (*GLS2)*, partially attenuated the causal relationship between glutamine and AD, and statistical colocalisation supported a shared signal among glutamine, blood *SPRYD4* expression levels, and AD. Further MR mediation suggested that glutamine mediated the association between *SPRYD4*gene expression and AD - consistent with a non-significant trend observed in our previous study using the Metabolon platform ^9^. These findings are supported by a recent GWAS study ^47^ reporting a novel AD SNP - rs2657879- in perfect LD with rs2939302 (R2=1, https://ldlink.nih.gov/) and which colocalised with *SPRYD4* expression in other tissues and cell types, like microglia, inhibitory neuron, spinal cord and macrophage. *SPRYD4* is located in the 12q13.3 locus, upstream of *GLS2,* and higher *SPRYD4* levels have been shown to inhibit cancer growth through apoptotic mechanisms ^56^. *GLS2*-encoding a liver-expressed glutaminase 2 required for hydrolysis of glutamine to glutamate- is the most biologically plausible candidate for the observed associations. Nevertheless, our analyses did not support a shared causal variant between *GLS2* and glutamine (PP4 = 3.97E-7) or between *GLS2* and AD (PP4=0.42), but indicated that *GLS2* and glutamine in this region are highly likely to be driven by independent causal variants (PP3=1), a finding in agreement with our previous work ^9^. Additionally, in the recently published AD GWAS^47^ only brain and not blood *GLS2* levels colocalised with AD ^47^. Future work investigating tissue-specific (brain vs blood) eQTLs, single-cell colocalization, and fine-mapping at 12q13.3 will be crucial to disentangle *SPRYD4* from *GLS2*, define the cell types mediating risk, and prioritize targets along the glutamine**-**glutamate axis.

Finally, our MR mediation analysis found that glutamine causally mediated the protective relationship between education and AD risk. Higher education is associated with greater cognitive and synaptic reserve. Glutamine plays a critical role in neurotransmitter cycling, serving as a precursor for glutamate and GABA, which are essential for synaptic transmission and plasticity. Higher education may therefore enhance glutamine metabolism to support these synaptic functions, thereby contributing to cognitive resilience. Taken together, our data implicate glutamine at the intersection of genetic susceptibility and modifiable risk, underscoring a plausible protective role in AD and the need for future mechanistic investigation.

BCAAs are essential amino acids that constitute approximately one-third of the total amino acid content in the human body ^57^. They are involved in critical cellular processes, including glutamate metabolism, protein synthesis, and energy production in the brain ^58^. Several observational studies—including ours—suggest that higher BCAA levels are associated with a reduced risk of dementia ^59^. However, experimental evidence from transgenic mouse models of AD has shown that lowering BCAA levels can alleviate AD-related pathology and cognitive decline. Furthermore, BCAAs have been implicated in promoting tau phosphorylation through the activation of the mTOR signalling pathway ^60^. Our MR study also identified leucine as a risk factor of AD in contrast to our observational analyses but consistent with findings from an independent MR study ^61^. When we investigated this relationship further using restricted cubic spline models, they revealed a non-linear (U-shaped or J-shaped) relationship between BCAAs (including leucine, valine and total BCAAs) and dementia subtypes (**Supplementary Figure 18**), suggesting that while moderate levels may be protective, excessively high levels could potentially be associated with increased dementia risk. These findings highlight the dual nature of BCAAs and emphasise the importance of maintaining a balanced BCAA level for brain health.

S_LDL_PL_pct and LDL_size were the only two metabolites demonstrating relatively consistent associations across individual Cox analyses, neuroimaging measures, and MR analyses, although the MR findings lacked robustness. S_LDL_PL_pct was associated with increased risk of dementia, reduced brain volume, and showed a causal relationship with higher AD risk. In contrast, LDL_size exhibited inverse associations. Interestingly, although S_LDL_PL_pct appears detrimental to brain health, it was linked to smaller WMHs. One possible explanation is that altered phospholipid content in small LDL particles reflects changes in lipid metabolism or inflammation ^62^ and impair neuronal or synaptic health ^63^, contributing to neurodegeneration and dementia risk independent of vascular injury. Supporting this, S_LDL_PL_pct was absent from the VaD metabolomic signature. Furthermore, S_LDL_PL_pct correlated with global grey matter and hippocampal volumes, but not with global white matter volume, suggesting a specific effect on neuronal and synaptic integrity. In contrast, LDL_size was primarily associated with VaD but not AD, linked to white matter volume and WMHs but not to global grey matter or hippocampal volume, indicating a predominantly vascular pathway in dementia risk. Taken together, these distinct patterns suggest that S_LDL_PL_pct influences dementia risk mainly through neurodegenerative mechanisms, whereas LDL_size could potentially contribute primarily via vascular pathways.

Beyond metabolomic signatures, we also found that a higher percentage of FC/TC in larger HDL particles is associated with an increased risk of dementia, whereas a higher percentage in smaller HDL particles appears protective—an observation not reported in previous studies. HDL particles absorb the active form of cholesterol-FC from peripheral tissues, initially forming small HDL particles that mature into larger HDL as FC is esterified into store form of cholesterol-CE within the particle, enabling cholesterol transport to the liver ^64^. However, an excess of FC in large HDL particles can lead to reverse diffusion of cholesterol back into peripheral cells ^65^ , potentially indicating HDL dysfunction. In contrast, smaller HDL particles are more actively involved in cholesterol uptake from tissues, so a higher FC ratio in these particles may reflect more efficient cholesterol clearance. Supporting our results, a recent study also observed high HDL FC content resulting in greater influx into macrophages ^66^, reinforcing the role of high HDL FC content in larger HDL as a marker of HDL dysfunction. Taken together, our findings suggest that the detailed lipid composition of specific lipoprotein subfractions may offer more precise insights into the mechanisms underlying dementia and serve as early dementia indicators.

Additionally, we also investigated the effect of sex and *APOE* ε4 on the relationship between metabolites and dementia, given their strong links with both metabolites and dementia ^67^. We found that protective effects of most metabolites against dementia were attenuated in *APOE* ε4 carriers or females. For example, PUFA ratio in all fatty acids including omega-3_pct and omega-6_pct appears to be less protective against dementia in females, consistent with our recent study showing that female participants with AD had reduced levels of highly unsaturated fatty acids□^7^ measured using LC-MS. Collectively, our results highlight that both *APOE* ε4 carrier status and sex shape how metabolites influence dementia risk, often reducing the protective benefits and altering harmful impacts, suggesting personalised approaches may be needed for prevention or treatment.

Our study has several limitations. First, the UK Biobank cohort is generally healthier than the general population, with participants more likely to be free of major diseases and adverse health behaviours ^68^. As a result, plasma metabolite levels in this cohort may be skewed toward a healthier profile, potentially underrepresenting individuals with metabolite imbalances that are detrimental to brain health. Despite this, we still observed U-shaped associations for several key metabolites, suggesting that even within a healthier population, higher or lower extremes of these metabolites may be associated with increased dementia risk. Second, we did not investigate the influence of lifestyle factors such as diet and physical activity on metabolite levels due to substantial missing data for these variables in the UK Biobank. Nevertheless, future studies in cohorts with more comprehensive lifestyle data are warranted to explore how these modifiable behaviours impact metabolomic profiles and dementia risk. Third, due to the current limitations of metabolite GWAS, we were unable to obtain summary statistics for all key metabolites—particularly for total BCAAs and metabolite ratios—which restricted our MR analyses. Finally, plasma metabolites may not fully reflect central nervous system metabolism or brain-specific pathological processes. Although peripheral metabolic changes can provide valuable insights, they may not directly capture the metabolic environment within the brain. Future research integrating CSF metabolomics or brain imaging-derived metabolic markers could help address this limitation.

## Conclusion

In this large-scale metabolomic study, integrating machine learning with causal inference approaches, we identified robust metabolic signatures predictive of dementia subtypes beyond conventional risk factors. Key metabolites—including LA_pct, glutamine, BCAAs, S_LDL_PL_pct, and LDL_size—were associated with structural brain changes, influenced the effects of established lifestyle risk factors (particularly obesity, with this effect strongly mediated by LA_pct), and showed evidence for causal involvement in dementia. These findings highlight metabolic dysregulation as an early, potentially modifiable contributor to dementia pathogenesis and suggest that blood metabolites may serve as valuable biomarkers for risk stratification, mechanistic insight, and targeted prevention strategies.

## Data availability

The data that support the findings of this study are available from the UK Biobank project site, subject to the registration and application process. Further details can be found at https://www.ukbiobank.ac.uk.

## Code availability

The code used in this study analysis code will be made available at https://github.com/Wolfson-PNU-QMUL/.

## Acknowledgements

We are grateful to the participants of the UK Biobank. The National Health Service (NHS) North West Centre for Research Ethics Committee approved the UK Biobank study (reference 11/NW/0382), which conforms to the provisions of the Declaration of Helsinki. We also thank the High Performance Computing cluster (Apocrita) at QMUL for facilitating these analyses, and all members of the Centre for Preventive Neurology for their ongoing support. The Centre for Preventive Neurology is supported by a grant from Barts Charity. This work was also supported by Alzheimer’s Research UK (SRF-2016A-3) and Parkinson’s UK (G2403). We acknowledge the participants and investigators of the FinnGen study (https://www.finngen.fi/en/access_results) for their contributions.

