## Supplementary Figures for "Circulating Metabolites are Linked to Dementia and Brain Imaging Phenotypes, and Mediate Modifiable Risk Pathways" (from another version of this preprint: version 1, 2025-11-02)

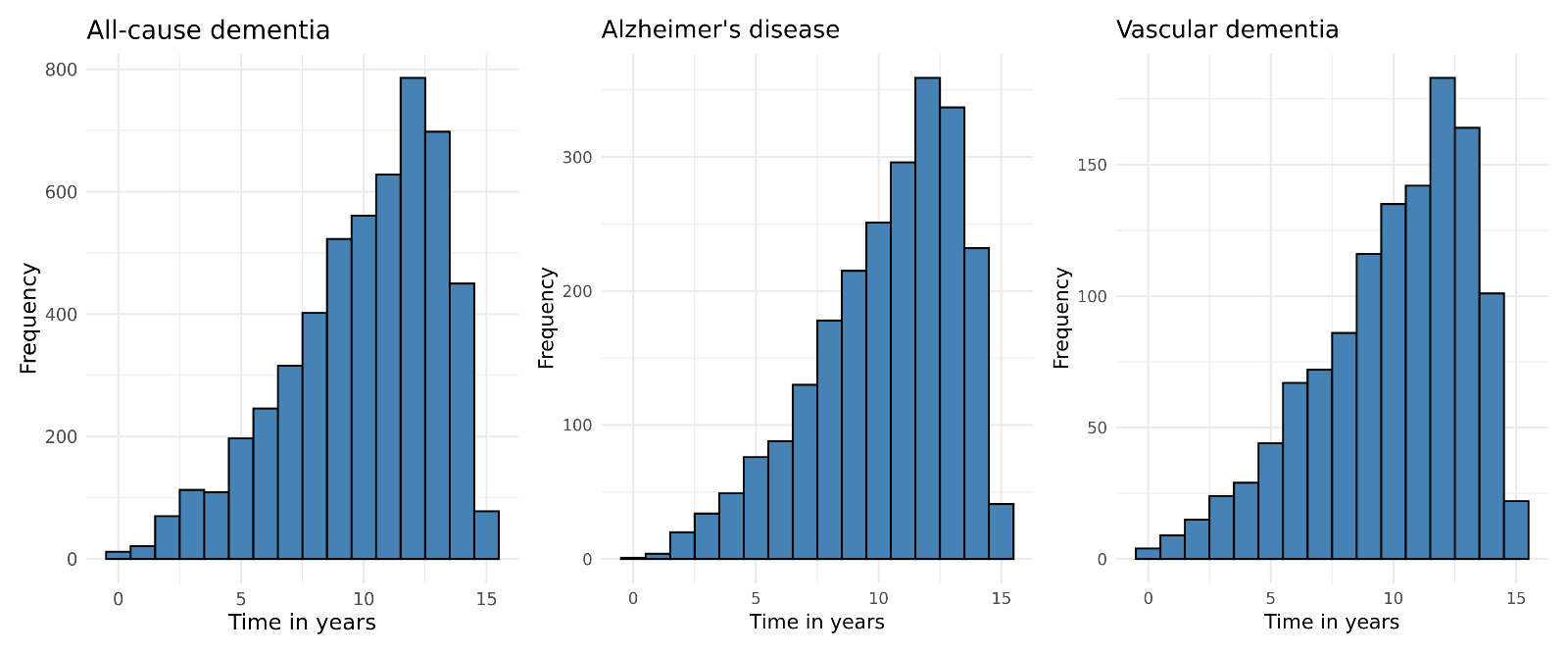
**Supplementary Fig. 1. Histogram of time-to-event of dementia subtypes:** all-cause dementia (ACD), Alzheimer’s disease (AD) and vascular dementia (VaD). The x-axis represents time in years (0–15), and the y-axis indicates the frequency of dementia cases.

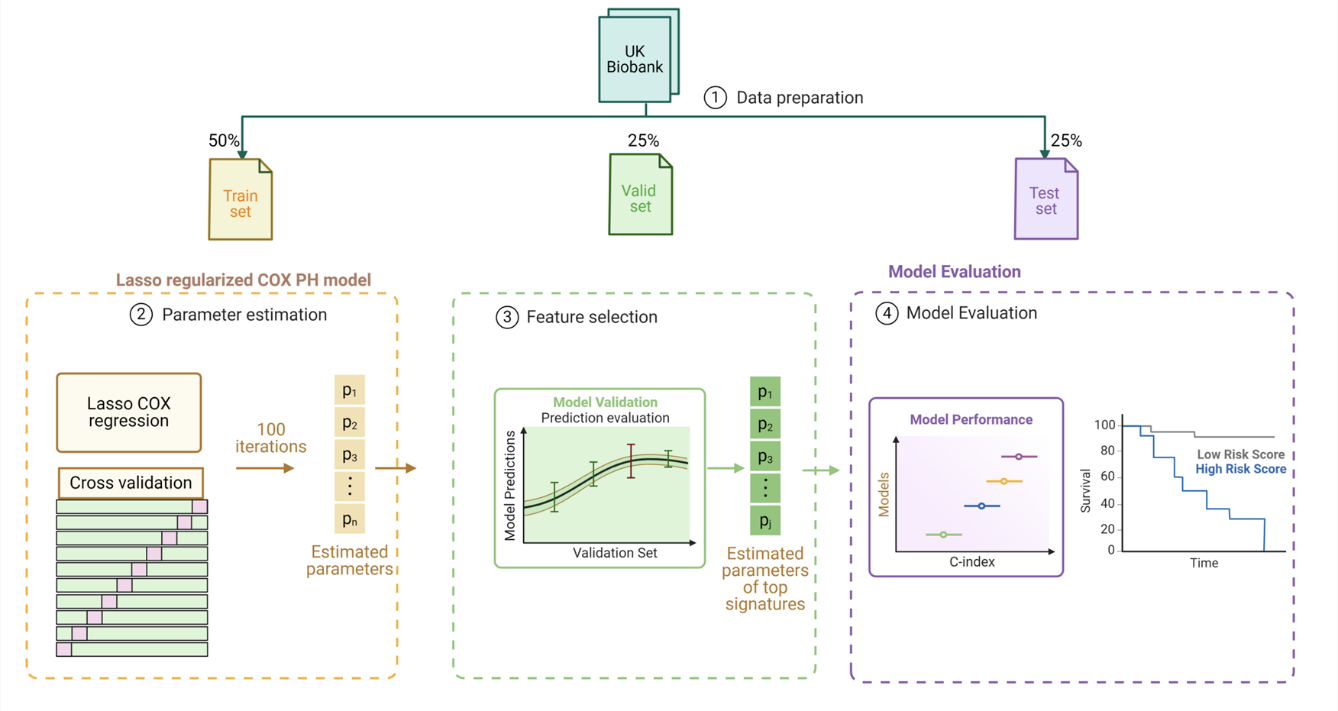
**Supplementary Fig. 2. Prediction model**

Predictive analysis was performed on the entire population without neuroimaging data. ①The data were split into training (50%), test (25%), and validation (25%) samples, with stratification to preserve the proportion of outcome classes across all splits. ② A lasso-regularised Cox regression model with 10-fold cross-validation (α = 1) was applied to the training set across 100 iterations. Metabolites were ranked by the mean absolute value of their regularised coefficients. ③ The C-index was evaluated in the validation set by sequentially adding top-ranked metabolites in groups of five. The final set of top metabolites was defined at the point where further additions no longer significantly improved the C-index (via bootstrapping). ④ Selected metabolites were used to construct a metabolomic risk score (MRS) in the test set to evaluate its predictive performance. Incremental improvements in the C-index were evaluated by adding the MetRS to three baseline models: model 1, model 2 and model 3. Model 1 adjusted for age, sex, ethnicity, Townsend index, centre, model 2 additionally adjusted for APOE ε4 carrier status, model 3 additionally adjusted for education level, obesity, smoking, alcohol, history of hypertension and diabetes. Finally, survival analyses were performed to assess the predictive performance of the MRS at 5-, 10-, and 15-year intervals.

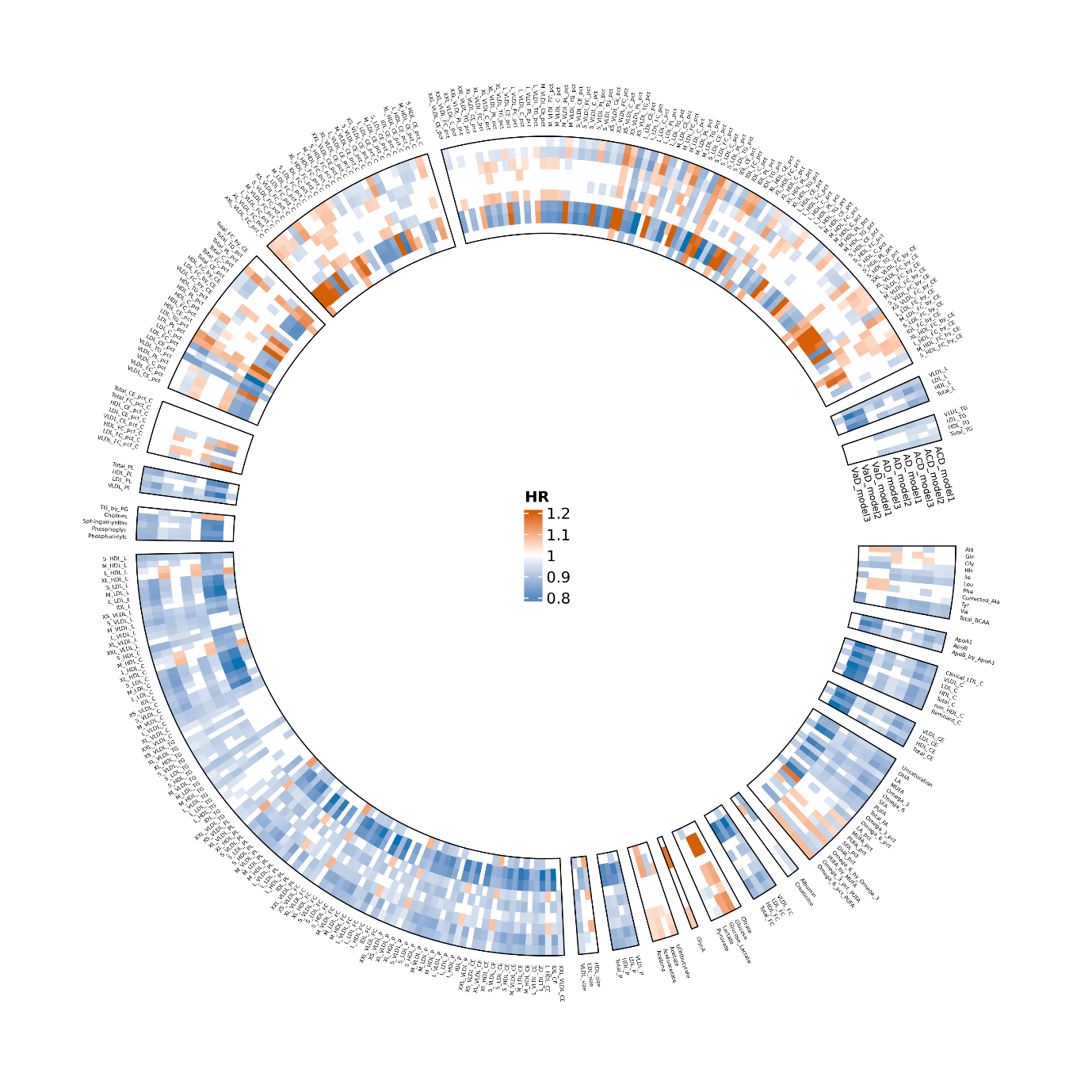
**Supplementary Fig 3. Circular plot showing associations between individual metabolites and dementia subtypes:** all-cause dementia (ACD), Alzheimer’s disease (AD), and vascular dementia (VaD).

Model 1 adjusted for age, sex, ethnicity, Townsend index, centre, model 2 additionally adjusted for *APOE* ε4 carrier status, model 3 additionally adjusted for education level, obesity, smoking, alcohol, history of hypertension and diabetes.

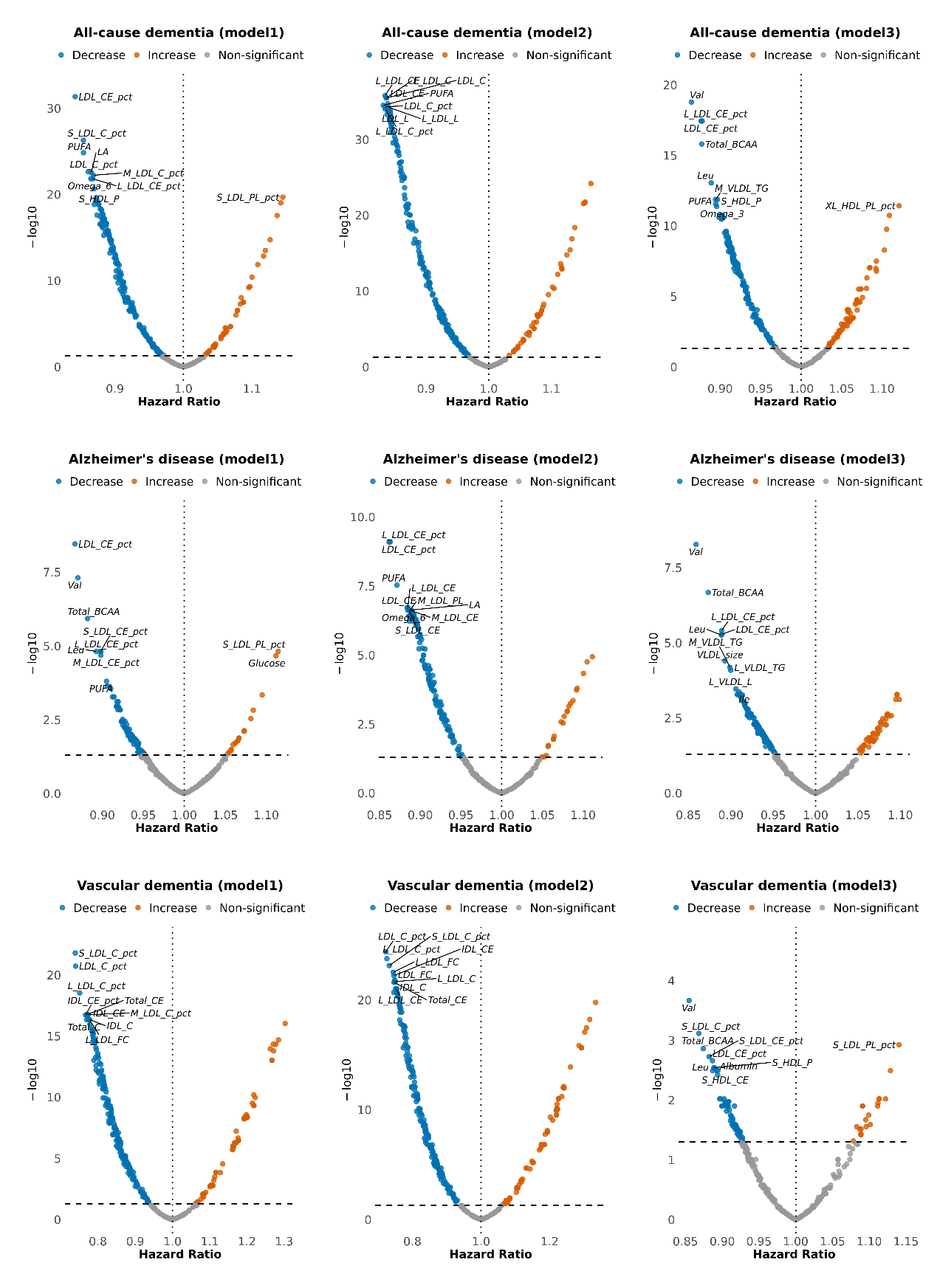

**Supplementary Fig.** **4. Volcano plots of the association between metabolites and risk of dementia subtypes:** All-cause dementia (ACD), Alzheimer’s Disease (AD), Vascular dementia (VaD). Model 1 adjusted for age, sex, ethnicity, Townsend index, centre, model 2 additionally adjusted for *APOE* ε4 carrier’s status, model 3 additionally adjusted for education level, obesity, smoking, alcohol, history of hypertension and diabetes.

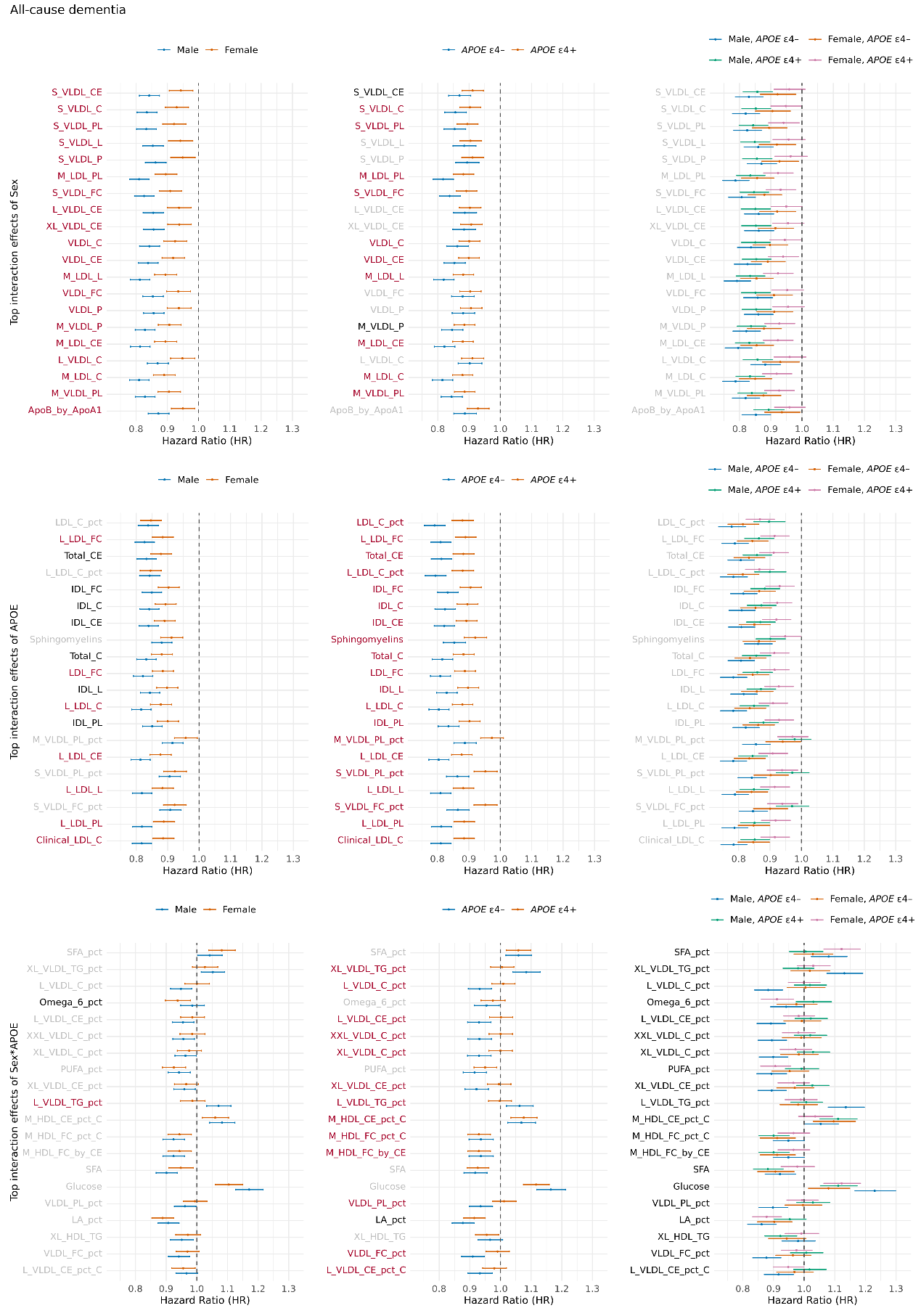

**Supplementary Fig. 5. Associations between metabolites and risk of all-cause dementia stratified by sex, *APOE* ε4 carrier status, and combined sex + *APOE* ε4 carrier status.** Each row presents the top 20 metabolite interactions for each stratification, alongside corresponding results for the same metabolites in other subgroup analyses. Metabolite labels are color-coded by interaction significance: grey for p > 0.05, black for p < 0.05 but FDR > 0.05, and red for FDR < 0.05.

Models were adjusted for age, sex, ethnicity, Townsend index, centre, and *APOE* ε4 carrier status (excluding the interacting variable for each interaction term).
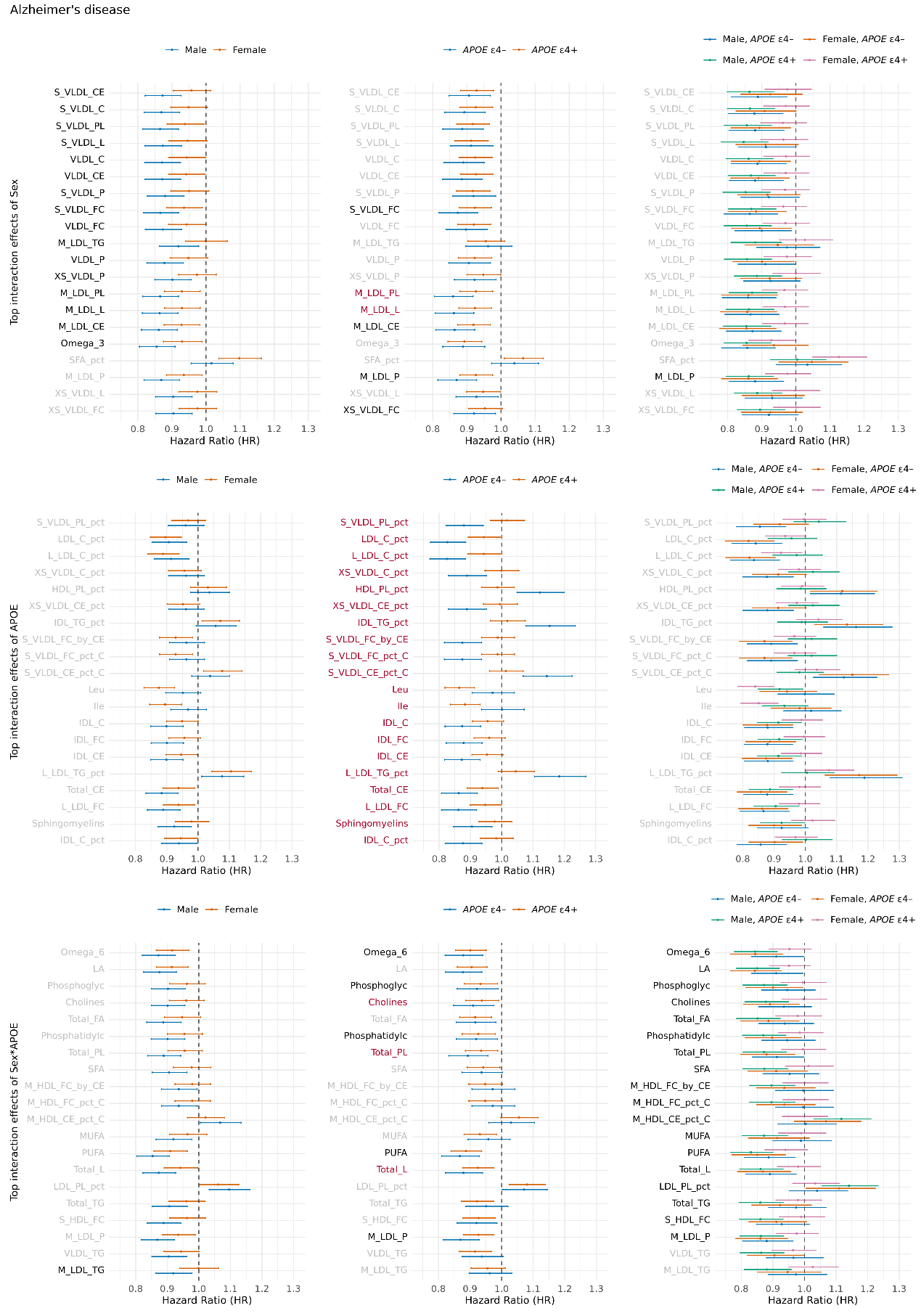
**Supplementary Fig. 6. Associations between metabolites and risk of Alzheimer’s disease stratified by sex, *APOE* ε4 carrier status, and combined sex + *APOE* ε4 carrier status.** Each row presents the top 20 metabolite interactions for each stratification, alongside corresponding results for the same metabolites in other subgroup analyses. Metabolite labels are color-coded by interaction significance: grey for p > 0.05, black for p < 0.05 but FDR > 0.05, and red for FDR < 0.05.

Models were adjusted for age, sex, ethnicity, Townsend index, centre, and *APOE* ε4 carrier status (excluding the interacting variable for each interaction term).

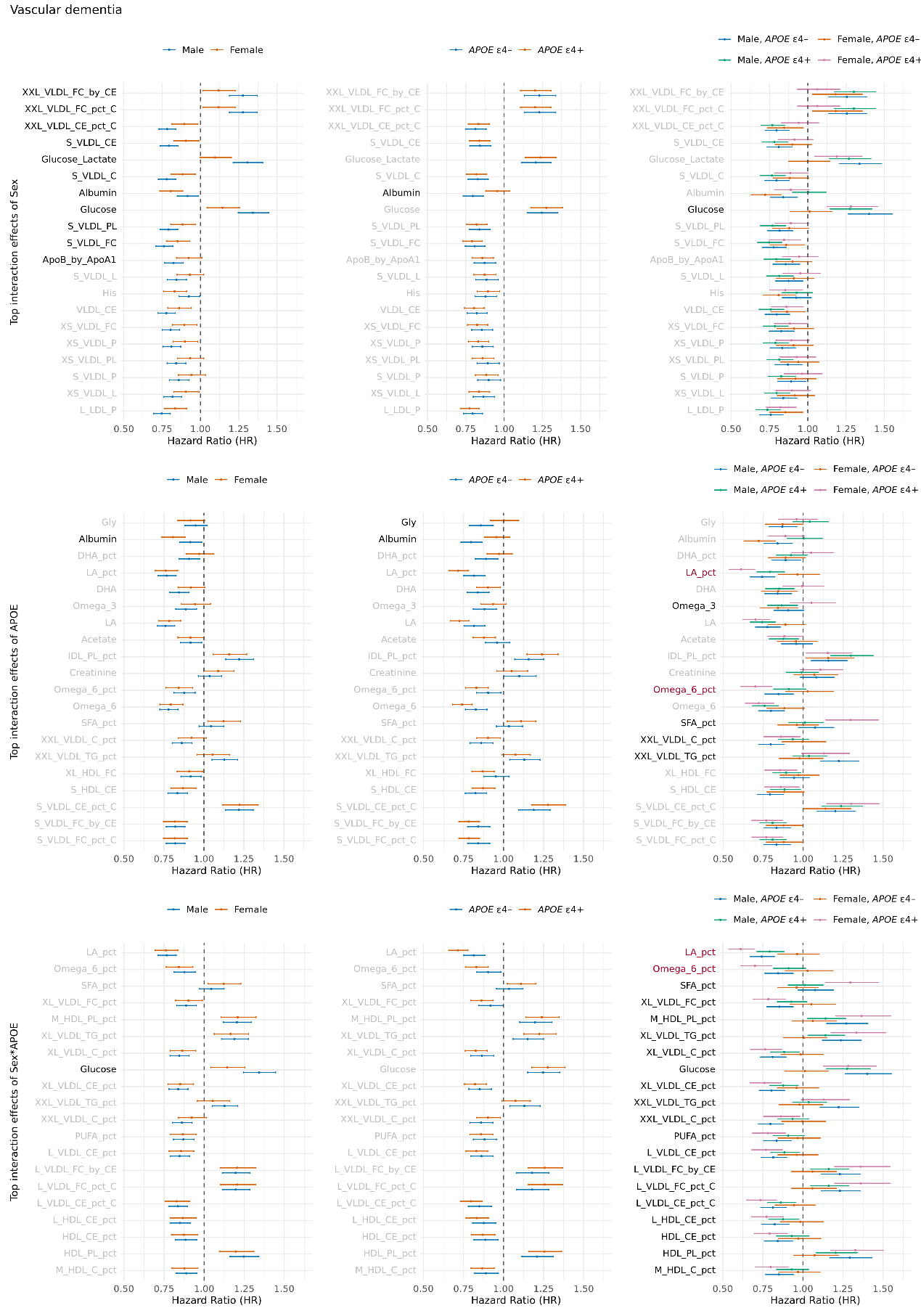
 **Supplementary Fig. 7. Associations between metabolites and risk of vascular dementia stratified by sex, *APOE* ε4 carrier status, and combined sex + *APOE* ε4 carrier status.** Each row presents the top 20 metabolite interactions for each stratification, alongside corresponding results for the same metabolites in other subgroup analyses. Metabolite labels are color-coded by interaction significance: grey for p > 0.05, black for p < 0.05 but FDR > 0.05, and red for FDR < 0.05.

Models were adjusted for age, sex, ethnicity, Townsend index, centre, and *APOE* ε4 carrier status (excluding the interacting variable for each interaction term).

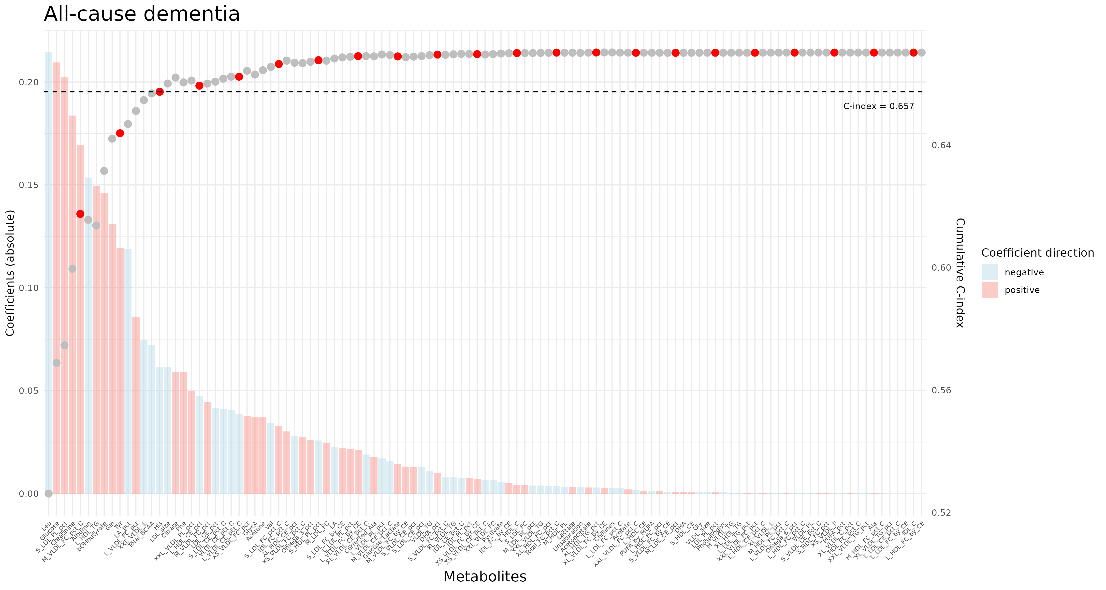

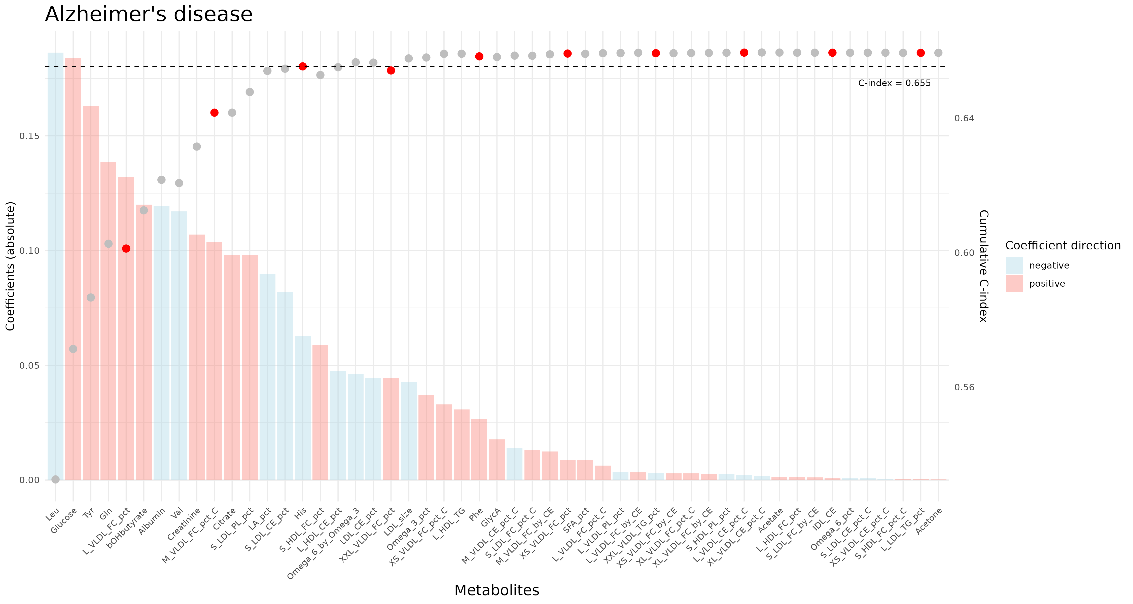

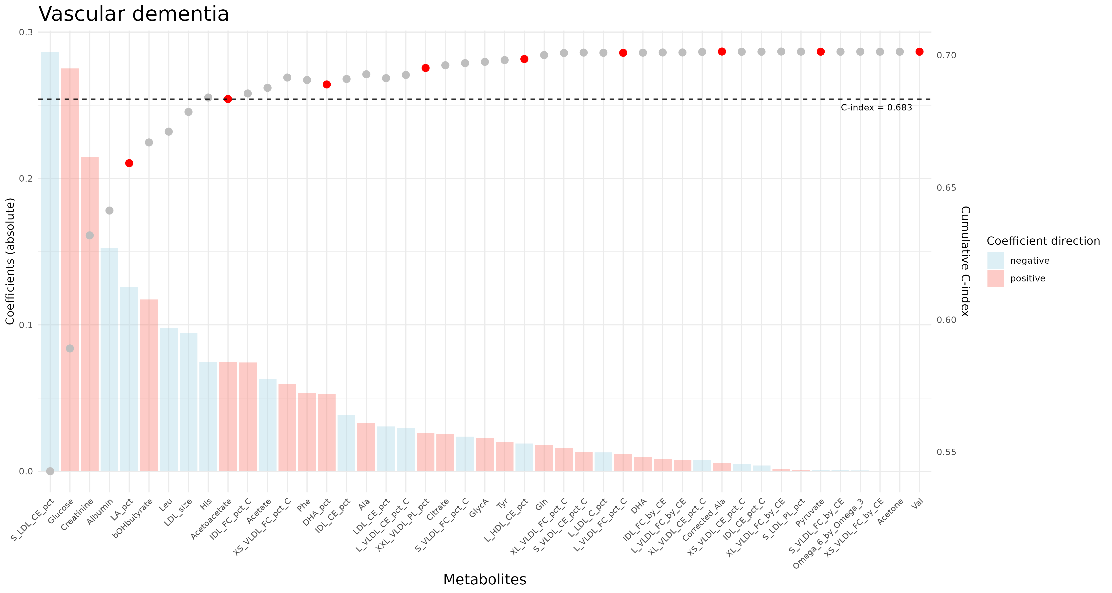

**Supplementary Fig. 8. The cumulative C-index of metabolites predicting dementia subtypes:** All-cause dementia (ACD), Alzheimer’s Disease (AD), Vascular dementia (VaD). Bar heights represent the absolute values of metabolite coefficients from a lasso regression model, with color indicating direction: lightblue for positive and salmon for negative associations. Overlaid points show scaled cumulative C-index values, with every fifth metabolite highlighted in red. The dashed horizontal line marks the accumulative C-index value of the final selected metabolite. Left y-axis shows coefficient magnitude; right y-axis shows cumulative C-index.

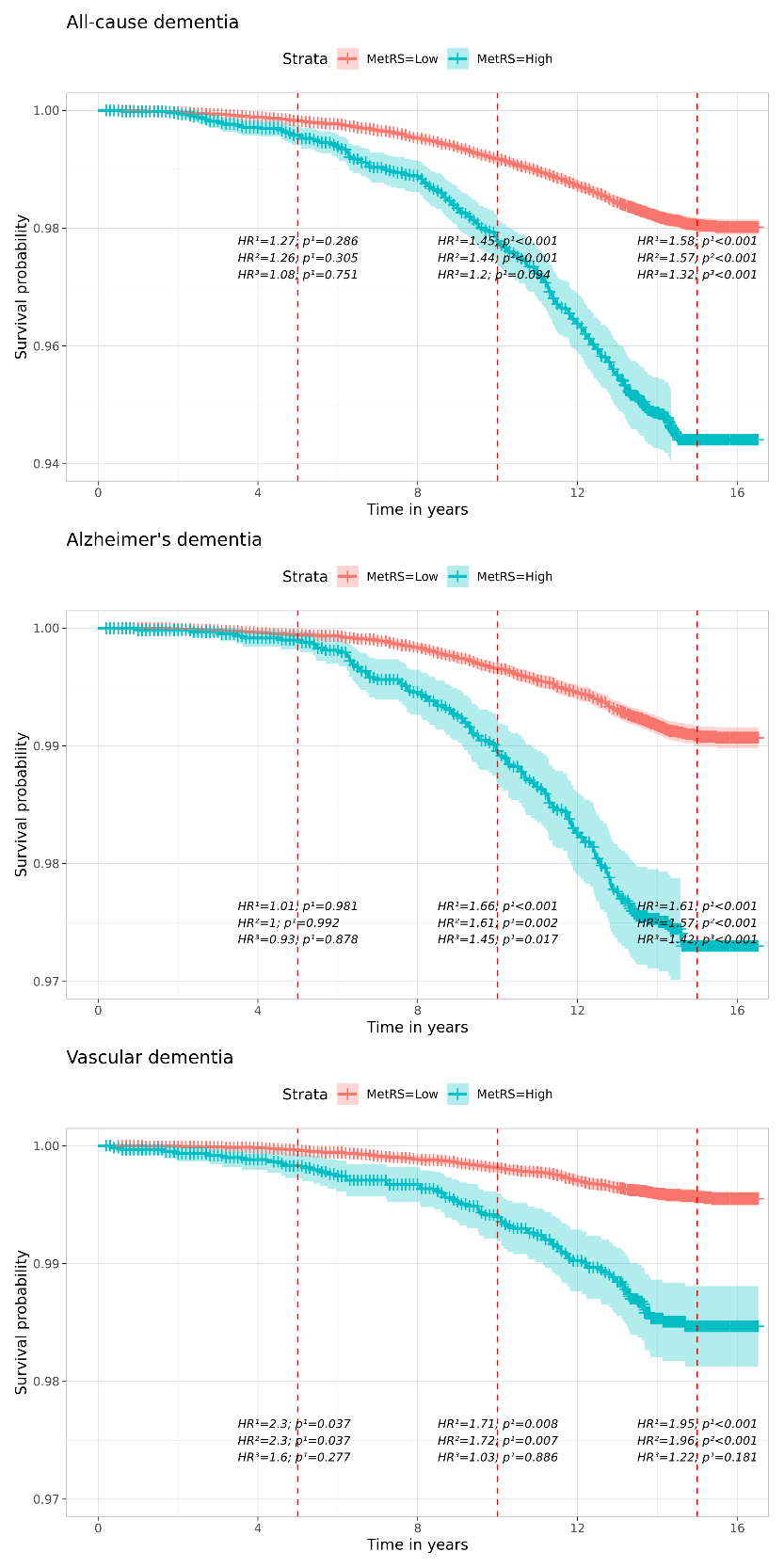

**Supplementary Fig. 9. Survival plots showing the predictive performance of metabolomic risk scores (MetRS) for dementia subtypes**—all-cause dementia (ACD), Alzheimer’s disease (AD), and vascular dementia (VaD)—at 5-year, 10-year, and 15-year follow-up intervals.

Model 1 adjusted for age, sex, ethnicity, Townsend index, centre, model 2 additionally adjusted for *APOE* ε4 carrier status, model 3 additionally adjusted for education level, obesity, smoking, alcohol, history of hypertension and diabetes.

^1^ model 1, ^2^ mode 2, ^3^ model 3.

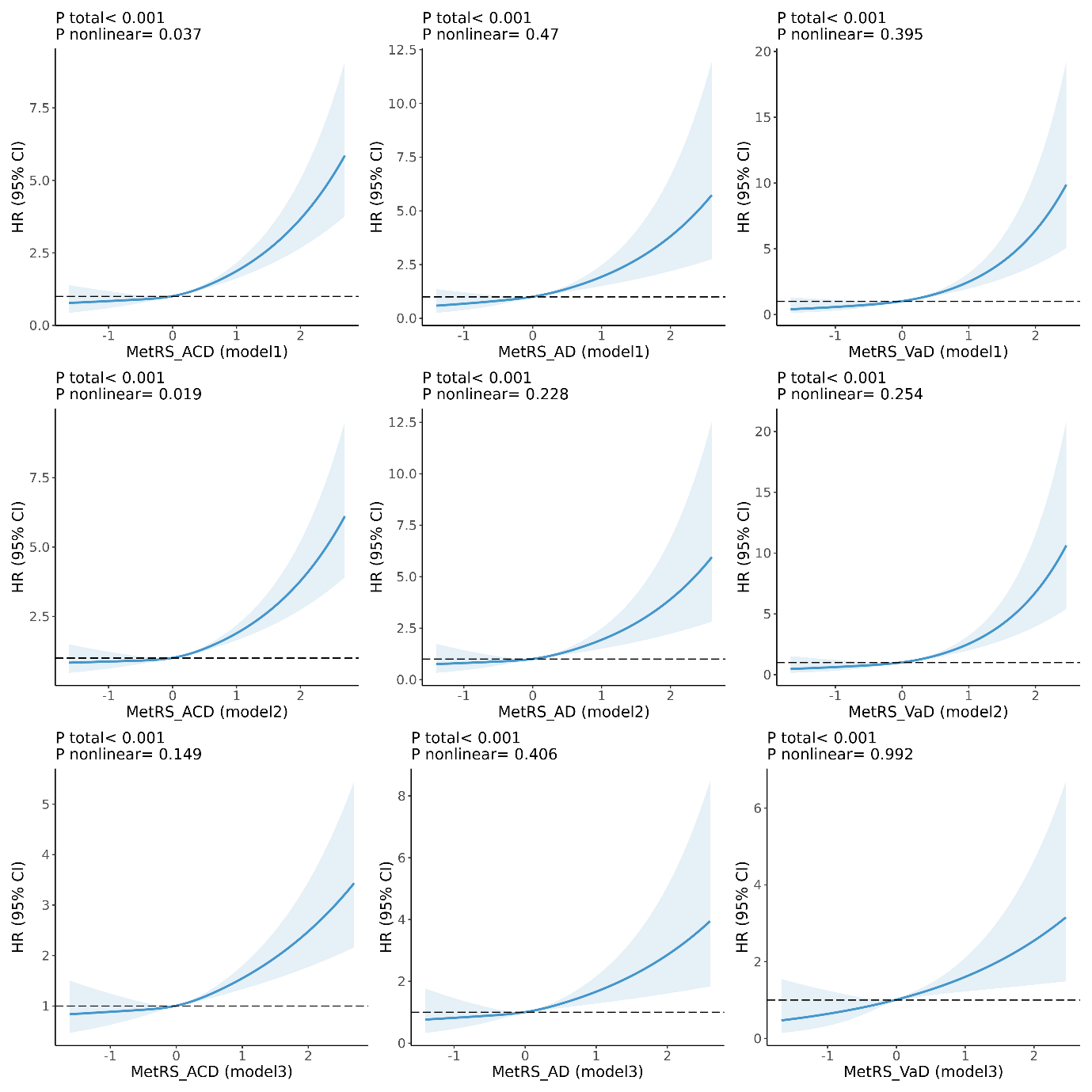

**Supplementary Fig 10. The dose-response curve of associations between metabolomics risk score (MetRS) and risk of dementia subtypes:** all-cause dementia (ACD), Alzheimer’s disease (AD), and vascular dementia (VaD).

Model 1 adjusted for age, sex, ethnicity, Townsend index, centre, model 2 additionally adjusted for *APOE* ε4 carrier status, model 3 additionally adjusted for education level, obesity, smoking, alcohol, history of hypertension and diabetes.

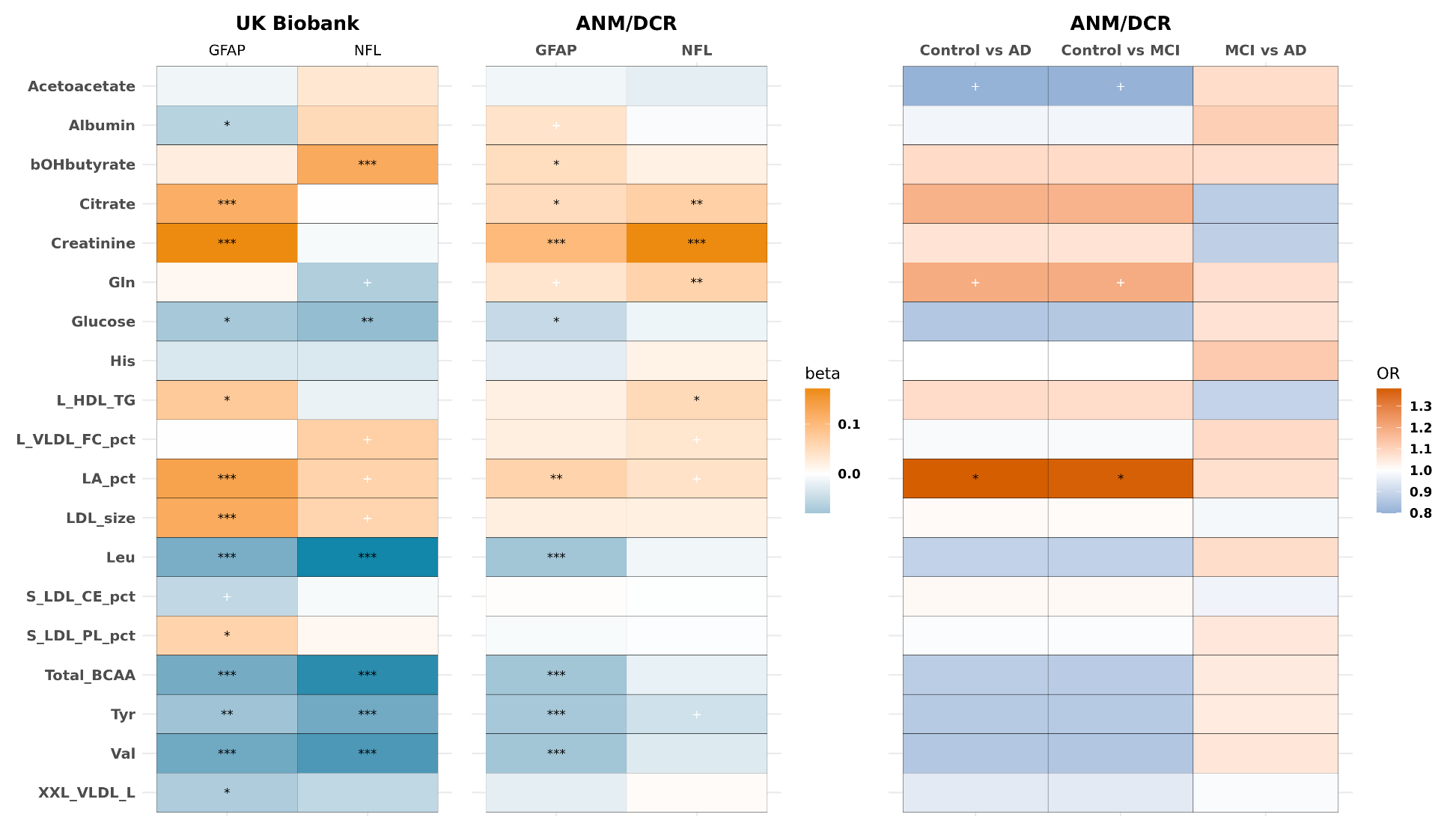

**Supplementary Fig 11. Associations between key metabolites and plasma Alzheimer’s disease biomarkers and case–control status in UK biobank and Addneuromed cohort.**

ANM/DCR, the AddNeuroMed/Dementia Case Register cohort.

Models were adjusted for age, sex, centre, and *APOE* ε4 carrier status.

+ p<0.05 and FDR>0.05, * FDR<0.05, ** FDR<0.01, *** FDR<0.001

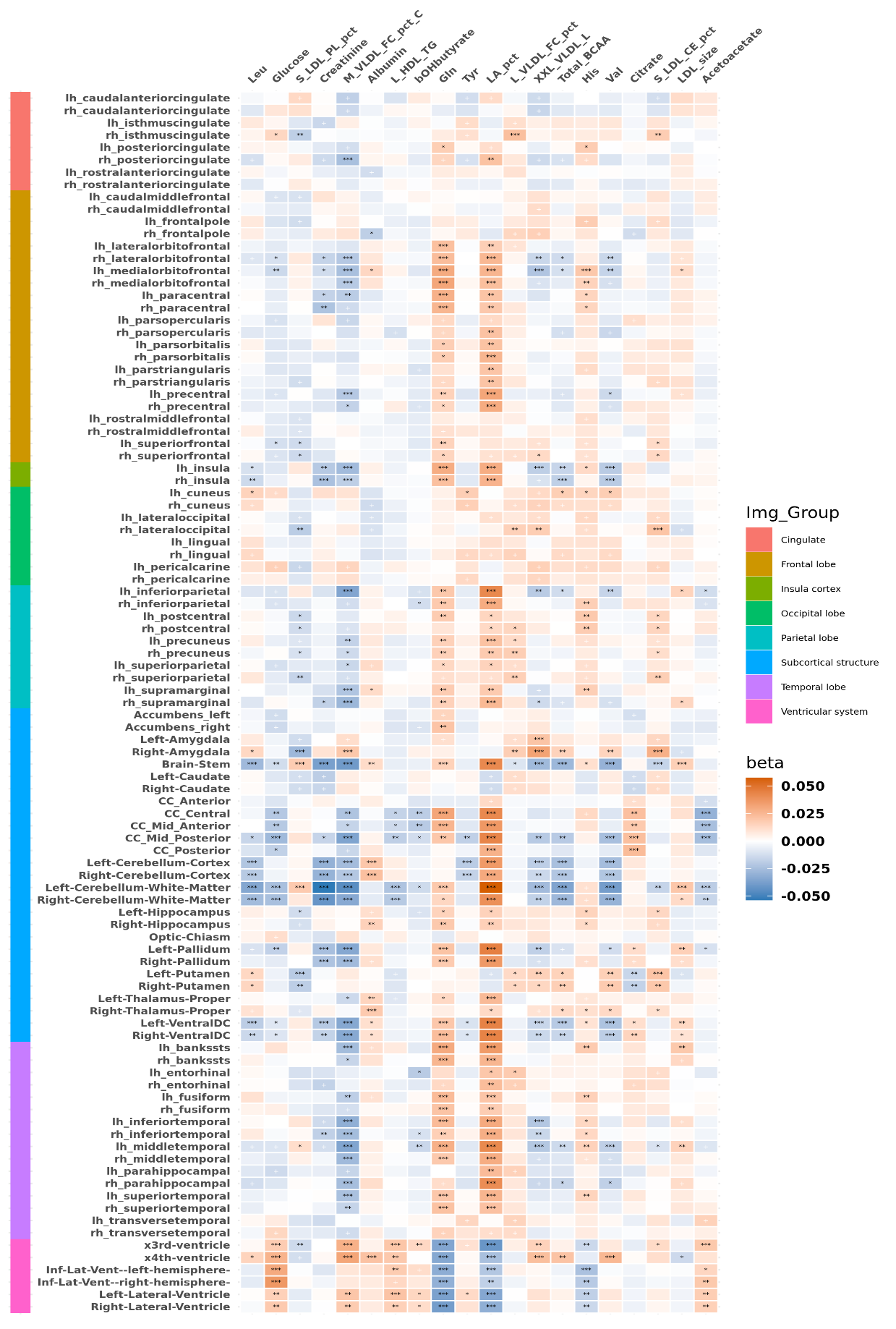

**Supplementary Fig 12. Associations between key metabolites and regional brain volume.**

Models were adjusted for age, sex, ethnicity, Townsend deprivation index, assessment centre, and *APOE* ε4 carrier status.

^+^ p<0.05 and FDR>0.05, * FDR<0.05, ** FDR<0.01, ***FDR<0.001

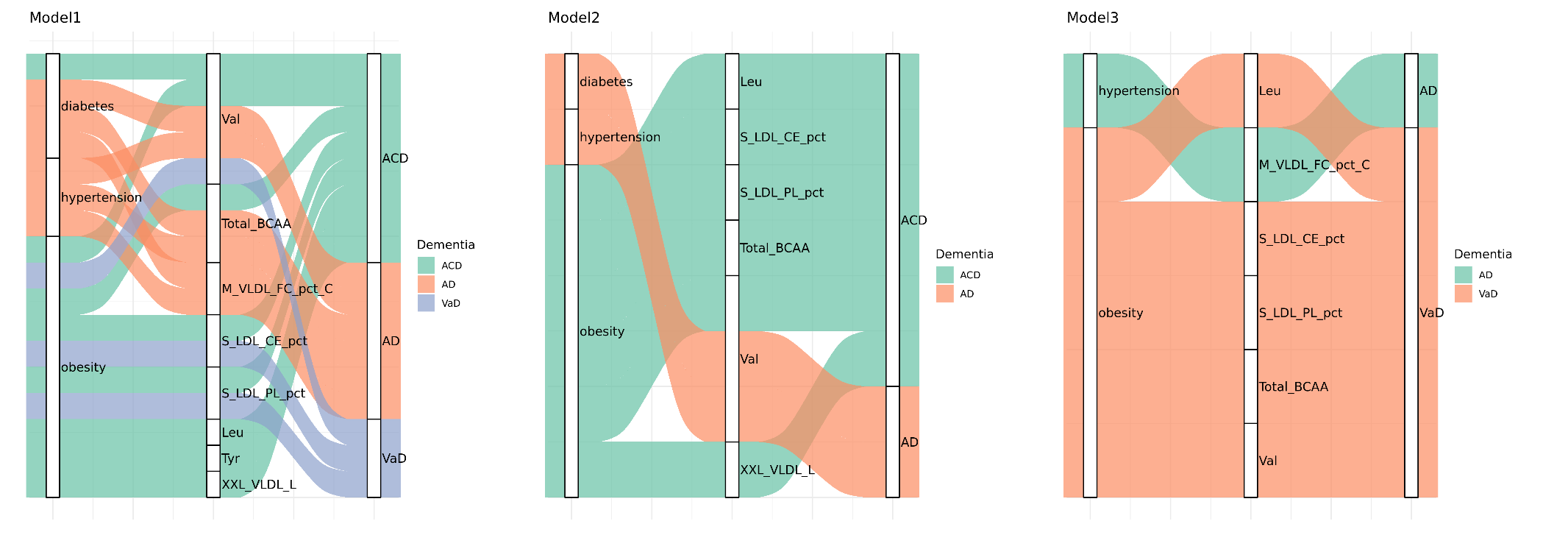
**Supplementary Fig 13. Sankey diagrams illustrating the suppressive effects of key metabolites in the relationship between risk factors and dementia subtypes:** all-cause dementia (ACD), Alzheimer’s disease (AD), and vascular dementia (VaD). A suppressive effect is defined as a proportion mediated (Prop.med) < 0, and only those with Prop.med < –10% are shown in diagrams. Each diagram displays pathways from modifiable risk factors (left panel), through key metabolites (middle panel), to dementia outcomes (right panel), across three models with increasing levels of covariate adjustment.

Model 1 adjusted for age, sex, ethnicity, Townsend index, centre, model 2 additionally adjusted for *APOE* ε4 carrier status, model 3 additionally adjusted for education level, obesity, smoking, alcohol, history of hypertension and diabetes.

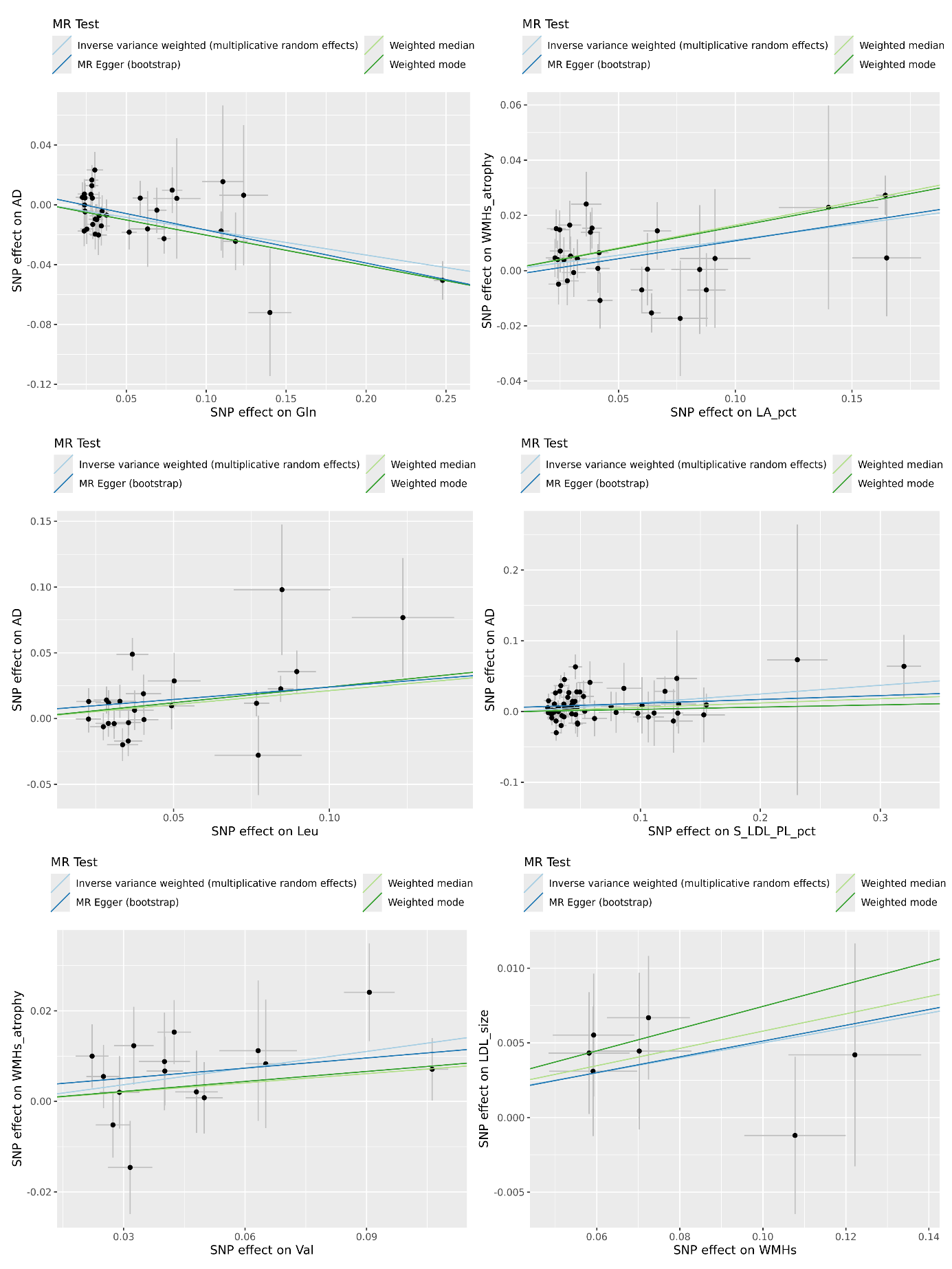

**Supplementary Fig 14. Scatter plots showing Mendelian Randomization (MR) analyses assessing causal relationships between metabolite levels and outcomes.** Only significant exposure–outcome pairs from bi-directional MR analyses based on inverse variance weighted (IVW) results with FDR < 0.1 are presented. Each plot displays SNP effects on the exposure (x-axis) versus SNP effects on the outcome (y-axis), with dotted cross-hairs indicating standard errors. Colored lines represent effect estimates from regression fits using different MR methods: IVW (multiplicative random effects), MR Egger (bootstrap), Weighted median, and Weighted mode.

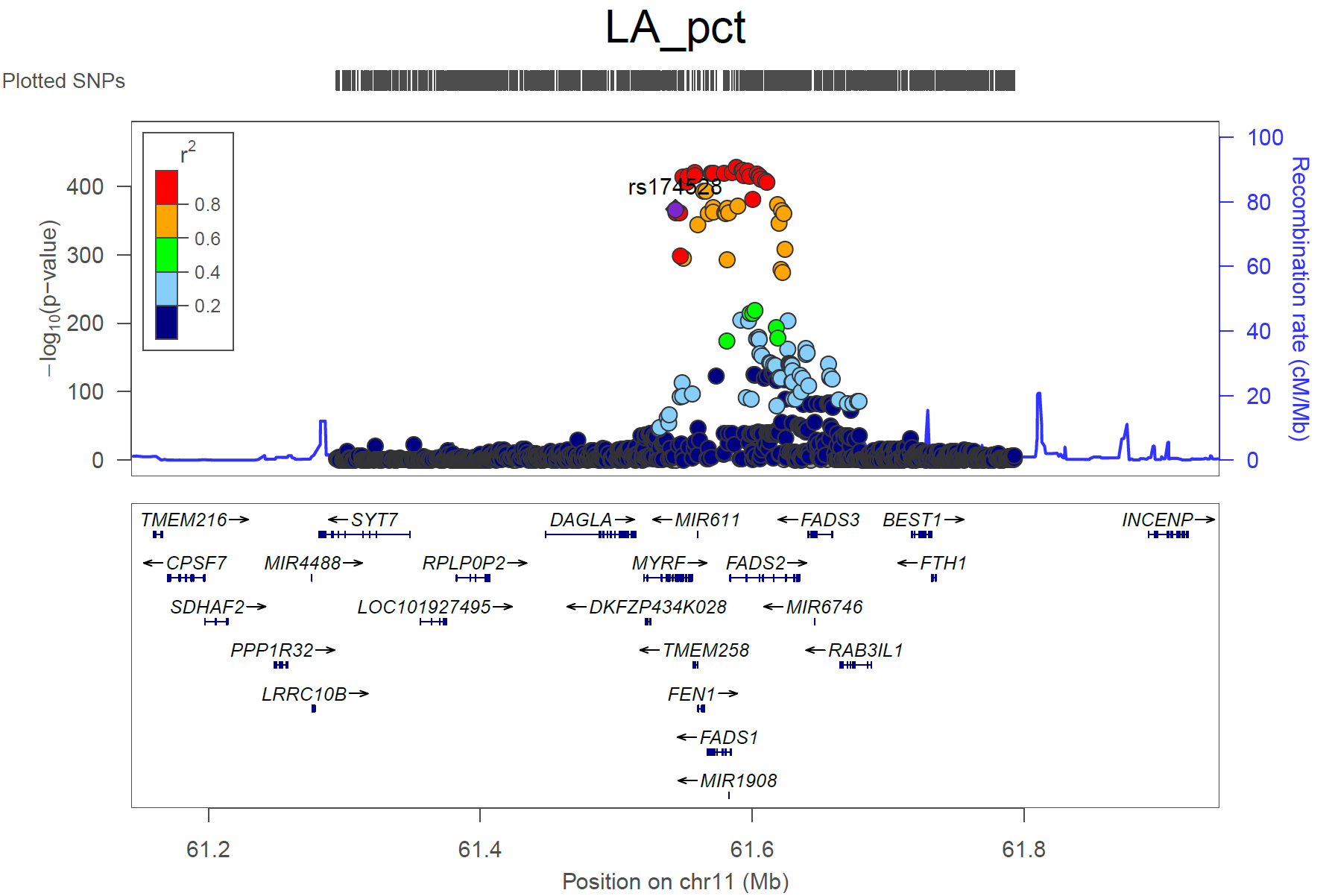

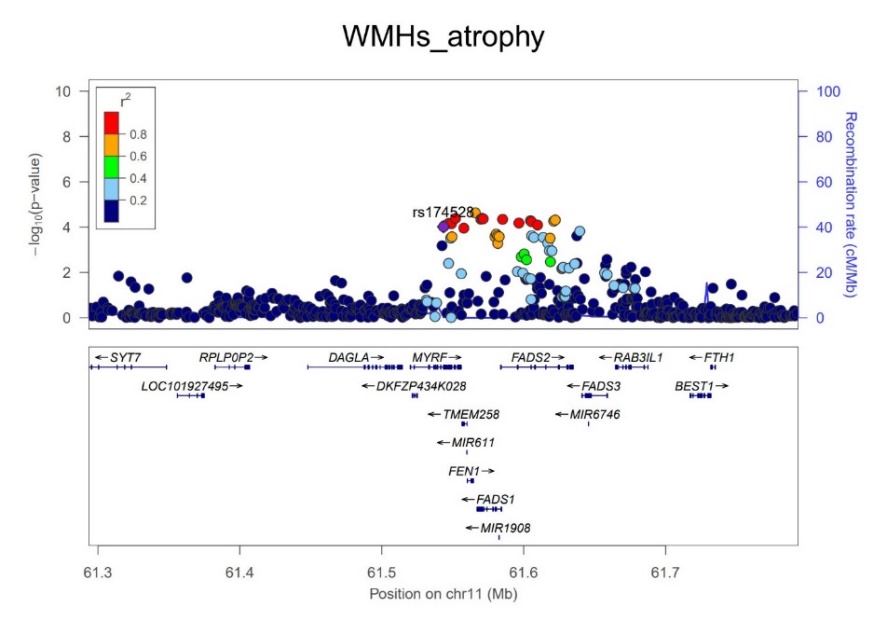

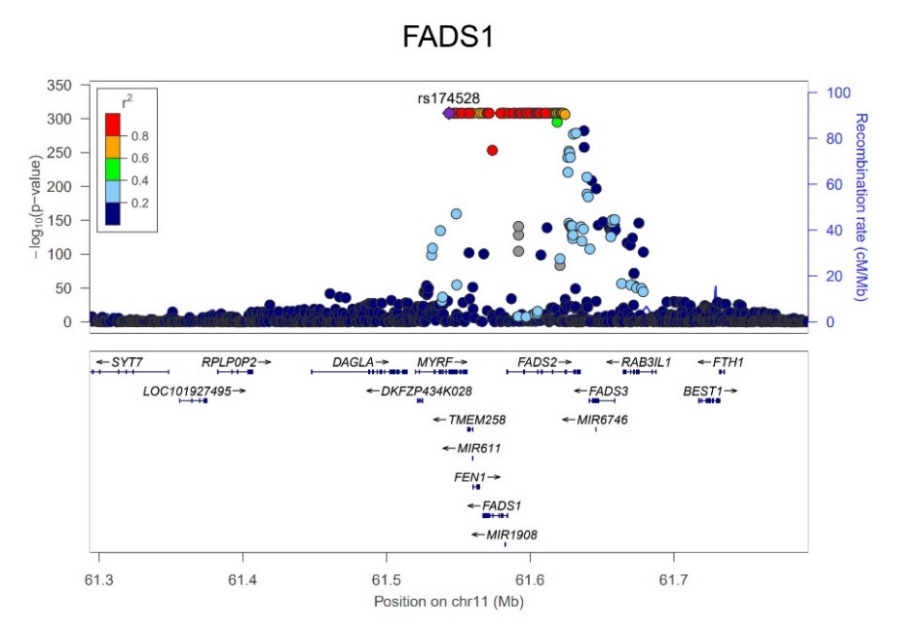

**Supplementary Fig 15. LocusZoom plots of the colocalising region Chr11:61293499-61793499 for LA_pct WMHs_atrophy and FADS1 gene.**

LA_pct, linoleic acid percentage to total fatty acids; WMHs_atrophy, white matter hyperintensities related brain atrophy.

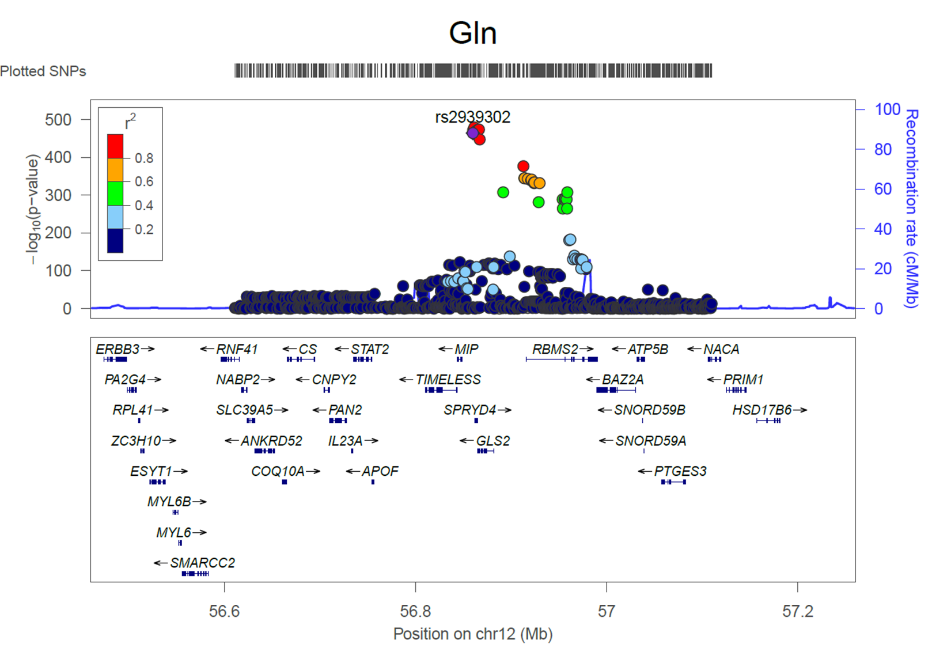

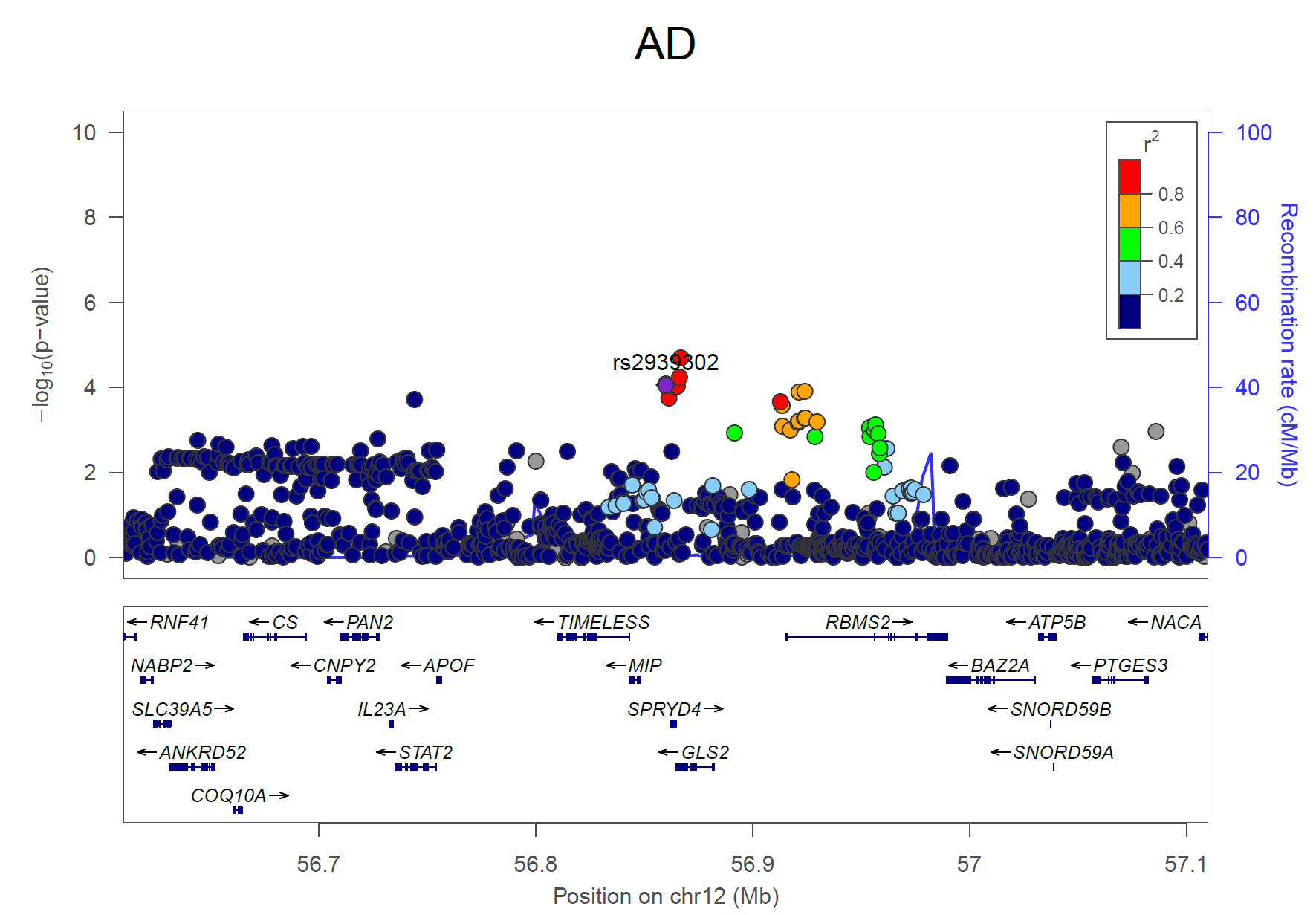

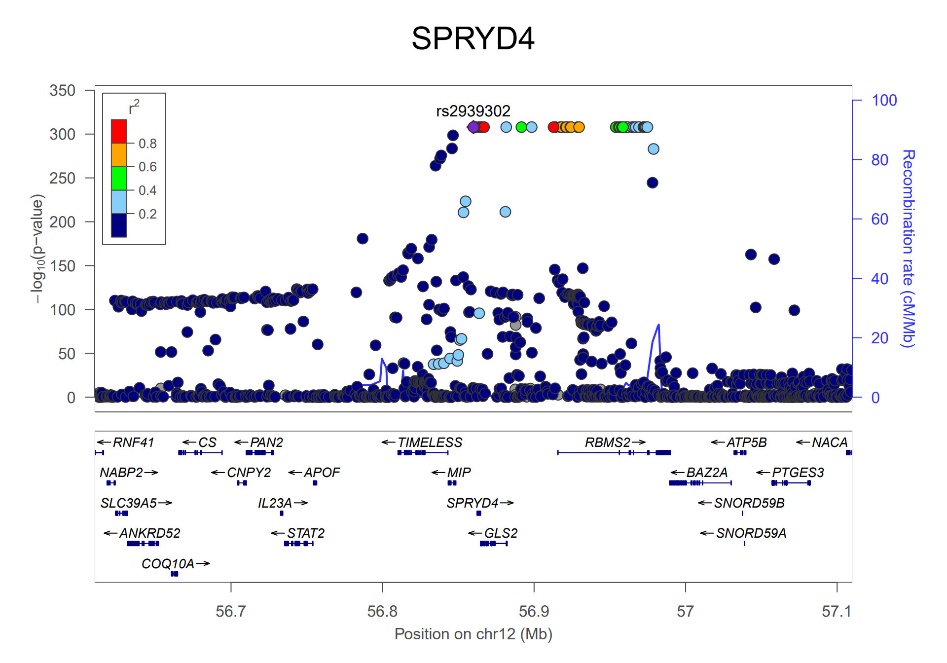

**Supplementary Fig 16. LocusZoom plots of the colocalising region Chr12:56610020-57110020 for glutamine, Alzheimer’s disease (AD) and SPRYD4 gene.**

SPRYD4, SPRY Domain Containing 4.

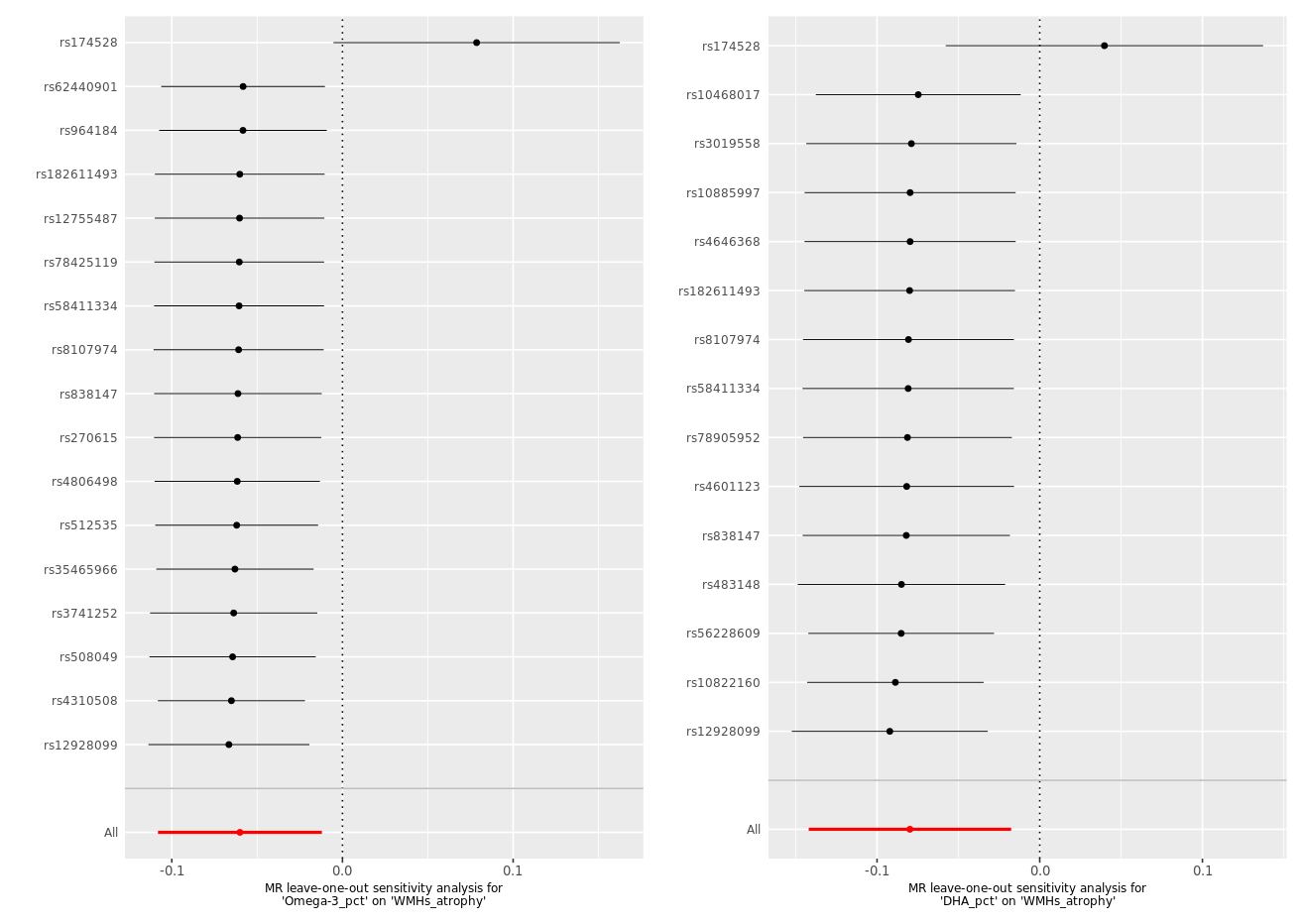

**Supplementary Fig 17. Leave-one-out sensitivity analyses for the association between omega-3 percentage in fatty acids and WMHs_atrophy, and between DHA percentage and WMHs_atrophy.**

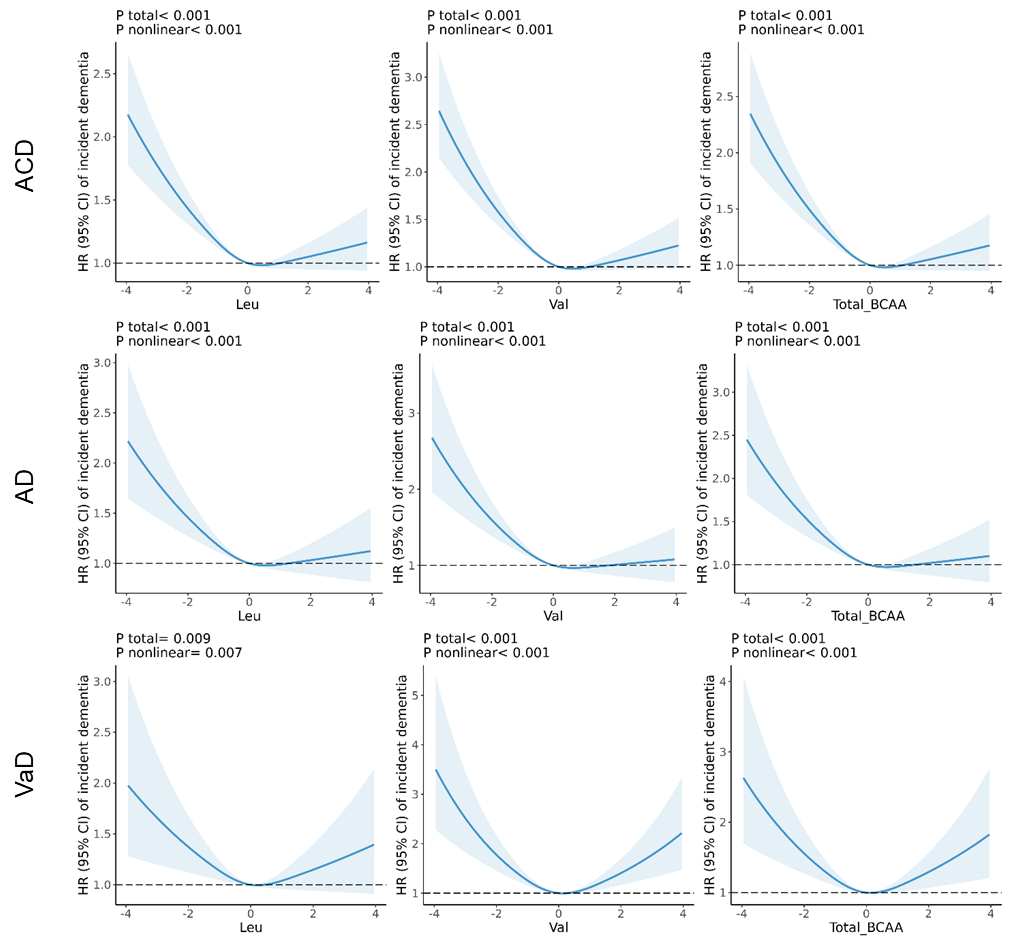

**Supplementary Fig 18. The dose-response curve of associations between branched-chain amino acids (BCAAs) and risk of dementia subtypes:** all-cause dementia (ACD), Alzheimer’s disease (AD), and vascular dementia (VaD).

Models were adjusted for age, sex, ethnicity, Townsend deprivation index, assessment centre, and APOE ε4 carrier status.

BCAAs include leucine (Leu), Valine (Val) and total branched-chain amino acids (BCAAs).
